# Dynamic Clinical States and Transitions During the First 72 Hours of Intensive Care After Acute Stroke

**DOI:** 10.64898/2026.08.30.26361738

**Authors:** Ping Lei, Yanyi Xu, Yuxia Zhang

## Abstract

**Background:** The condition of a patient with acute stroke often changes within hours of ICU admission. Prognostic work here targets fixed endpoints predicted from admission data, and trajectory phenotyping assigns one label per patient. We used longitudinal ICU data to identify interpretable dynamic clinical states, characterize transitions between them, and relate the current state to later events.

**Methods:** Retrospective cohort study of 6368 adults with acute stroke in MIMIC-IV v3.1. The first 72 h were divided into twelve 6-hour windows, and a hidden Markov model was fitted to 21 neurological, physiological and organ-support variables. State number was chosen against criteria fixed before fitting: statistical fit, restart stability, state occupancy and clinical interpretability. Generalized estimating equations related the current state to new mechanical ventilation and vasopressor use within 12 h, and to ICU death within 72 h. Eleven sensitivity analyses assessed the robustness of the state solution.

**Results:** Four states were selected: neurologically preserved-low support, neurological impairment- low support, impairment-renal dysfunction and impairment-respiratory support (63.3%, 7.8%, 11.8% and 17.1% of windows). Within 72 h, 40.3% of patients changed state at least once, and transitions ran in both directions rather than along a single severity gradient. States were identified without outcome data, yet ICU mortality by last state ranged from 2.9% to 43.9%. Adjusted for age, sex, subtype and Charlson index, the current state remained associated with organ-support escalation and death. State prevalence differed by at most 1.1 percentage points between training and test sets, and 10 of 11 sensitivity analyses gave a stable four-state solution (ARI 0.754–0.955).

**Conclusions:** The early ICU course of acute stroke can be represented as movement among a small number of clinically interpretable states. The representation was reproducible in a held-out set and across admission eras, but requires validation in an independent database before any clinical use.

## 1. Introduction

A patient admitted to the ICU after acute stroke is not in a fixed condition. Level of consciousness, respiratory and circulatory support, renal function and treatment intensity can all change within a few hours. The same patient may deteriorate and then improve, or move back and forth between very different clinical situations. At the bedside, what matters is not only whether the patient will ultimately die, but where the patient is now, what has changed over the past few hours, and what is most likely to happen next.

Prognostic research in stroke critical care has largely been organized around fixed endpoints — in-hospital death, the need for mechanical ventilation, functional outcome — predicted from variables recorded in the first hours after admission[1]. Models of this kind estimate how likely a given endpoint is, but they say little about the course itself[2]. A clinician may know that a patient’s risk of death is high and still not know what clinical situation that patient is currently in, or how that situation is changing.

Trajectory analysis has more recently been used to describe longitudinal change in critically ill patients, including in ischemic stroke, where distinct vital sign trajectory phenotypes have been identified and linked to treatment response[3]. This work uses the longitudinal data, but it still resolves to one label per patient: a phenotype is assigned from the pattern observed over an interval, and the patient carries it for the whole period. The same design has recently been applied to the Glasgow Coma Scale in a multicentre cohort of 7876 patients with stroke, where four latent trajectory classes carried prognostic information beyond baseline variables[4]. In both cases the longitudinal data are used to place the patient in one class for the whole period. A patient who requires mechanical ventilation on admission, is successfully weaned, and then remains on minimal support has passed through a transition that a single label, however well derived, cannot express.

These three approaches can be set out as a progression in what is being represented. Conventional prognostic modeling maps baseline features to an outcome. Trajectory phenotyping maps longitudinal data to one phenotype per patient. The present study maps longitudinal data to a state that the patient occupies at each point in time, to the transitions between those states, and from there to the next clinical event. The unit of representation is not the patient but the patient-window, and the object of interest is not the label but the movement between labels. What distinguishes this from conventional patient-level trajectory phenotyping is that the state assignment is allowed to change from one window to the next.

We therefore used multidimensional longitudinal data from the first 72 h of ICU care in acute stroke to identify dynamic clinical states and to describe how patients move between them. We then examined whether the current state carries information about subsequent organ-support escalation and death. Finally, as an exploratory secondary analysis, we compared the distribution of states across the three major stroke subtypes.

## 2. Methods

### Data source and study design

This was a retrospective cohort study using MIMIC-IV v3.1[5], a de-identified critical care database from a single academic medical center in Boston, United States. No new data were collected. Reporting follows the STROBE statement for observational studies[6].

### Study population

We included adults whose hospital admission carried a code for acute ischemic stroke, intracerebral hemorrhage or subarachnoid hemorrhage among the first three diagnosis positions, and who were admitted to an ICU during that hospitalization[7]. Admissions also coded for traumatic brain injury were excluded to reduce misclassification of traumatic hemorrhage. Only the first ICU stay of the qualifying stroke admission was retained for each patient.

| Subtype | ICD-9 | ICD-10 |
| --- | --- | --- |
| Acute ischemic stroke (AIS) | 433.x1, 434.x1 | I63.x |
| Intracerebral hemorrhage (ICH) | 431 | I61.x |
| Subarachnoid hemorrhage (SAH) | 430 | I60.x |

The primary analysis required an ICU stay of at least 12 h, so that every patient contributed at least two 6-hour windows and one state transition could therefore be observed. The threshold was relaxed from the 24 h originally planned mainly to avoid systematically excluding the most severely ill patients, who die within the first ICU day; the 24 h threshold was retained as a sensitivity analysis.

### Observation windows and definition of a clinical state

Observation began at ICU admission and continued to 72 h, or until ICU discharge or death, whichever came first. The first 72 h were divided into twelve consecutive, non-overlapping 6-hour windows. Windows after discharge or death were treated as right-censored.

We use *clinical state* for the multidimensional condition of a patient within a single 6-hour window, *state transition* for a change of state between two adjacent windows, and *clinical trajectory* for the ordered sequence of states a patient passes through during the observation period.

A clinical state as defined here is not a purely physiological construct. It is characterized jointly by neurological responsiveness, physiological measurements, and the organ support the patient is concurrently receiving. Treatment is included deliberately: what a clinician recognizes at the bedside as a patient’s current situation includes the support that patient is on, and a state defined on physiology alone would place a ventilated patient and a self-ventilating patient with identical gas exchange in the same state. Because this makes treatment part of the definition, a prespecified sensitivity analysis refits the model with all four organ-support variables removed, to establish whether the structure survives without them.

### Variables

Twenty-one variables were used to identify states: 17 continuous and 4 binary organ-support variables. Variable selection balanced coverage of the physiological domains routinely assessed in critical care against the recording density needed to support analysis at 6-hour resolution in MIMIC-IV[8].

| Domain | Variables |
| --- | --- |
| Neurological | GCS eye score, GCS motor score |
| Respiratory | Respiratory rate, peripheral oxygen saturation |
| Circulatory | Heart rate, systolic blood pressure, mean arterial pressure |
| Renal-metabolic | Creatinine, blood urea nitrogen, sodium, potassium, glucose, 6-hour urine output |
| Hematological | White cell count, hemoglobin, platelet count |
| Systemic | Temperature |
| Organ support (binary) | Invasive mechanical ventilation, continuous renal replacement therapy, continuous vasopressor infusion, continuous sedative infusion |

Continuous variables were summarized as the last recorded value within each window, which stays closest to the patient’s condition at the end of that window; binary treatment variables were coded positive whenever the treatment overlapped the window in time. Lactate, pH and four arterial blood gas variables exceeded a prespecified 70% window-level missingness threshold and were not included.

The GCS verbal score was not included in the primary model. Verbal response cannot be assessed directly in an intubated patient: in this cohort the verbal score was unavailable in 29.0% of windows, and in 80.2% of those windows the patient was receiving mechanical ventilation. Assigning all unassessable records either the lowest score or the median would systematically overstate or understate neurological impairment. The primary analysis therefore describes neurological responsiveness using the eye and motor scores and their sum (GCS-EM, range 2–10)[9]; by comparison, GCS-EM itself was missing in only 5.3% of windows. Three alternative treatments of the verbal score were carried as sensitivity analyses.

### Data handling

Data cleaning removed only physiologically impossible values. Values that were clinically extreme but plausible were retained after inspection of the source records (Supplementary Methods S1).

Before any imputation parameter was computed, patients were split 70/30 into a training and a held-out test set, stratified by stroke subtype, with all windows from a given patient kept in the same set. Continuous variables were first carried forward within patient for a limited period — up to 6 h for vital signs and neurological variables, up to 12 h for laboratory values — and remaining missing values were filled with the training-set median (Supplementary Methods S5). Variables were then standardized to z-scores using training-set means and standard deviations. All parameters estimated in the training set were frozen before being applied to the test set.

### Identification and naming of dynamic states

A dynamic clinical state cannot be observed directly; it can only be inferred from the physiological and treatment measurements recorded at the same time, and states at adjacent time points are typically related to one another. We therefore fitted a hidden Markov model in the training set[10,11]. The model summarizes the multidimensional observations of each window as a latent state and describes change over time through a transition matrix. The 17 continuous variables were modeled with diagonal-covariance normal emissions and the 4 binary variables with categorical emissions; parameters were estimated by Baum-Welch expectation-maximization, and the most likely state for each window was obtained by Viterbi decoding.

Models with 3 to 6 states were fitted and compared. The dimensions on which candidate models would be evaluated were fixed in the statistical analysis plan before any model was fitted (Supplementary Methods S2), and were: (i) statistical fit, by the Bayesian information criterion and log-likelihood; (ii) the share of patients and of windows falling in each state; (iii) whether any state was very small; (iv) whether the states differed clinically in a substantive way; (v) whether results were stable across random initializations; and (vi) whether clinicians could reasonably interpret the states.

The analysis plan also stated in advance that the number of states would not be chosen on the Bayesian information criterion alone, because in latent class trajectory analysis small changes in model specification can materially alter both the recovered classes and their apparent clinical associations. No single criterion was therefore treated as decisive, and the final choice was a judgment across all six dimensions rather than the output of a fixed decision rule. Supplementary Methods S2 reports each criterion for every candidate model and states how the criteria were weighed.

Robust standardization using medians and interquartile ranges, which had also been prespecified, degenerated to a single-state solution under this emission model and was therefore not used for the primary analysis (Supplementary Methods S3).

The silhouette coefficient is reported for the selected model as a descriptive measure of static geometric separation in the raw feature space. It was not one of the state-selection criteria and was not used to choose K: silhouette treats each window as an independent point and is therefore blind to the transition structure that distinguishes a hidden Markov state from a cluster. It is computed on the full training set and shown in Supplementary Fig. S3.

State indices carry no clinical meaning, so each state was given a descriptive name based on its most prominent neurological, organ-function and support characteristics. Naming used no outcome information and was adjudicated by a critical care nurse scientist (Y.Z., RN, PhD) who was blinded to all outcome data at the time of review (Supplementary Methods S4). These names summarize the dominant features of a state; they are not criteria that every window assigned to that state must satisfy.

### Transitions, outcomes and subtype analysis

Transition probabilities were estimated both from the fitted transition matrix and empirically from the Viterbi-decoded state sequences. State duration was measured as the number of consecutive 6-hour windows spent in the same state. When describing common trajectories, consecutive self-transitions were collapsed so that the sequence reflects the changes a patient actually underwent.

Mortality enters this study in two distinct ways, which answer different questions and should not be read as one result. The first is descriptive: patients are grouped by the state they occupied in their last observed window, and ICU mortality is reported within each group. This describes the outcome distribution associated with each state at the patient level and involves no model. The second is an association analysis: each 6-hour window is treated as a discrete-time hazard interval, and the state occupied in a given window is related to death occurring during the first 72 h, adjusted for baseline characteristics. This is a window-level, time-varying analysis. The first is not an estimate of risk conferred by a state, and the second is not an estimate of overall ICU mortality.

Three window-level outcomes were prespecified: new mechanical ventilation within the next 12 h, new vasopressor use within the next 12 h, and ICU death within the first 72 h. The first two analyses were restricted to windows in which the patient was not already receiving that form of support. Associations between the current state and subsequent events were estimated with generalized estimating equations, adjusted for age, sex, stroke subtype and the Charlson comorbidity index[12–14], with clustering by patient. For mortality, each 6-hour window was treated as a discrete-time hazard interval and death was counted as an event only in the window in which it occurred. Odds ratios were reported only for states contributing at least 10 outcome events; this rule was fixed before the results were examined. Landmark survival analyses were performed at 24 h and 48 h to reduce the immortal-time bias that arises when a time-varying state is used as a baseline grouping[15,16].

Window-level state distributions were compared across the three subtypes with a chi-square test, and ICU mortality was compared across subtypes within the same last observed state.

### Sensitivity analyses

Robustness of the state solution was examined in two groups of analyses, distinguished by when they were specified rather than by what they found.

*Prespecified sensitivity analyses*, named in the study design document before any model was fitted, were: removing all treatment variables; adding laboratory missingness indicators; refitting with routinely available variables only, implemented as removing all laboratory variables; removing forward filling altogether; a complete-case analysis restricted to windows in which every continuous variable was measured; excluding patients with high sedation depth; restoring the 24 h stay threshold; restricting the cohort to AIS; using 12-hour windows; and using within-window means instead of last values.

*Additional robustness analyses* were specified during execution, in response to problems that emerged while the analysis was carried out, and are labelled as such throughout: three alternative treatments of the GCS verbal score, which the design document flagged as requiring special handling without naming a solution; robust standardization, adopted as the intended primary scaling after the plan was written; and refitting with uncapped age, which is reported as a change in the estimated odds ratios rather than as an agreement statistic and therefore does not appear in the sensitivity table.

Except where no comparable state structure could be formed, each was compared with the primary model using the adjusted Rand index[18]. The design document listed further sensitivity analyses that were not performed; these are itemized in the accompanying analysis plan.

### Software

Analyses were performed in Python 3.9 using pandas, scikit-learn, statsmodels, pomegranate 1.1.2 and plotly[17]. Analysis code is available from the corresponding author upon reasonable request. MIMIC-IV data are governed by their data use agreement and cannot be redistributed.

Generative AI tools (OpenAI ChatGPT; Anthropic Claude and Claude Code) were used to help implement the analysis code, generate the figures, and draft manuscript text. They were not used to make study design decisions, to define variables or the cohort, to choose analytic specifications, or to interpret the results. Every numerical result reported here was re-derived from the underlying data and checked against the analysis outputs by the corresponding author, and the clinical characterization of the identified states was adjudicated independently by a co-author blinded to outcome data. The authors reviewed and edited all AI-assisted output and take full responsibility for the content.

## 3. Results

### Study population

The final cohort comprised 6368 patients: 3461 with AIS (54.3%), 2022 with ICH (31.8%) and 859 with SAH (13.5%); a further 26 patients (0.4%) carried codes for both ICH and SAH and were handled separately. Median age was 69 years [IQR 57–80], 50.0% were women, and 714 patients (11.2%) died in the ICU. Patients with SAH were younger and had a lower comorbidity burden but a longer ICU stay than the other two subtypes (Table 1).

**Table 1.**
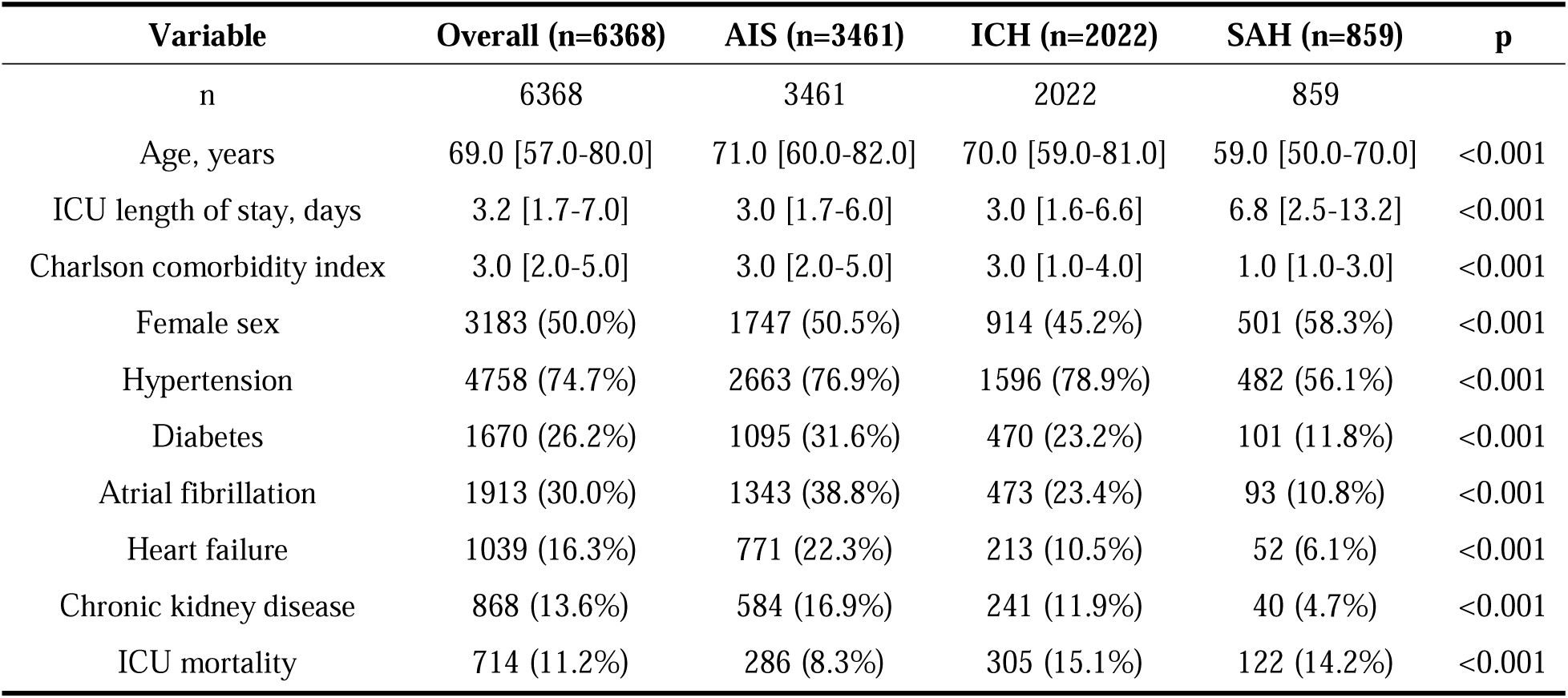
Baseline characteristics, overall and by stroke subtype.

**Table 2.** Candidate state-count (K) model comparison.

| **Candidate K** | **Log-likelihood** | **BIC** | **Min. state occupancy (%)** | **Restart stability (ARI)** |
| --- | --- | --- | --- | --- |
| 3 | -721,152 | 1,443,606 | 13.5 | 0.983 |
| 4 | -717,281 | 1,436,345 | 7.9 | 0.995 |
| 5 | -676,187 | 1,354,657 | 9.1 | not estimable |

*K=5 and K=6 excluded: each converged in only 1 of 50 restart attempts, so restart stability cannot be assessed.*

After stratification by stroke subtype, patients were split into a training set of 4457 and a test set of 1911, contributing 43261 and 18457 six-hour windows respectively, for a total of 61718 windows.

### Four dynamic clinical states

Across the six evaluation dimensions fixed before fitting, the four-state solution gave the most reproducible and clinically distinguishable representation (Supplementary Methods S2). The five-state model had a lower BIC, but converged in only 1 of 50 restarts and its additional state did not separate a clinically distinct group.

The clearest difference among the four states is in neurological responsiveness. Among the three states with neurological impairment, renal function and organ-support requirements then separate patients further. In neurologically preserved-low support, GCS-EM is near-complete and organ-support requirements are low. In neurological impairment-low support, neurological responsiveness is reduced but organ support is still rarely needed. Neurological impairment-renal dysfunction is additionally marked by clear renal abnormality, and neurological impairment-respiratory support by high mechanical ventilation and sedation requirements. (see Table 3)

**Table 3.** Clinical profile of each dynamic state (median [IQR] unless stated)

| State | Window share (%) | Patients ever in state (%) | ICU mortality if last state (%) | GCS-EM | Heart rate | SBP | MAP | Creatinine | BUN | Mech vent (%) | CRRT (%) | Vasopressor (%) | Sedative (%) |
| --- | --- | --- | --- | --- | --- | --- | --- | --- | --- | --- | --- | --- | --- |
| Neurologically preserved-low support | 63.3 | 80.7 | 2.9 | 10 [10-10] | 76 [66-87] | 132 [118-146] | 88 [78-98] | 0.90 [0.70-0.90] | 16 [12-17] | 6.9 | 0.0 | 4.2 | 7.1 |
| Neurological impairment-low support | 7.8 | 18.5 | 3.8 | 8 [7-9] | 82 [70-93] | 138 [124-153] | 89 [79-100] | 0.90 [0.80-1.10] | 16 [14-22] | 4.3 | 0.0 | 2.0 | 6.8 |
| Neurological impairment-renal dysfunction | 11.8 | 18.6 | 28.0 | 7 [5-9] | 84 [72-96] | 125 [108-141] | 80 [70-90] | 1.90 [1.20-3.00] | 38 [26-52] | 56.8 | 1.4 | 25.4 | 39.4 |
| Neurological impairment-respiratory support | 17.1 | 31.2 | 43.9 | 6 [4-7] | 82 [71-95] | 129 [114-141] | 83 [74-92] | 0.90 [0.70-0.90] | 16 [12-17] | 78.0 | 0.0 | 20.0 | 58.2 |

At the level of individual measurements, the states differ mainly in neurological responsiveness, mechanical ventilation requirement and renal function. Median GCS-EM across the four states was 10, 8, 7 and 6; mechanical ventilation was in use in 6.9%, 4.3%, 56.8% and 78.0% of windows; and median creatinine was 0.90, 0.90, 1.90 and 0.90 mg/dL. Routine vital signs such as heart rate and blood pressure differed comparatively little between states.

No mortality or other outcome information was used to identify the states. Even so, when patients were grouped by their last observed state, ICU mortality differed markedly across the four groups, from 2.9% to 43.9%. This is the descriptive, patient-level comparison defined in the Methods: it characterizes the outcome distribution associated with each state, and is a different quantity from the window-level association with death reported in Section 3.4. It does not imply that the four states form a fixed severity ladder.

The four states overlap substantially in the raw feature space: the mean silhouette coefficient over the full training set was 0.017 (Supplementary Fig. S3). This is reported as a descriptive measure of static geometric separation and was not among the criteria used to select the model. Silhouette evaluates each window as an independent point, so it cannot see the transition structure that distinguishes these states from clusters, and physiological change in the ICU is in any case continuous rather than grouped into separable regions. The evidence bearing on whether the structure is meaningful is presented in the sections that follow: the clinical profile of each state, the regularity of transitions between them, reproducibility in the held-out test set, and association with subsequent clinical events.

### Patients move continually between states

Within the first 72 h, 2567 patients (40.3%) changed state at least once: 1185 occupied 2 states, 761 occupied 3, and 621 occupied 4 or more. The remaining 59.7% stayed in a single state throughout, most of them in neurologically preserved-low support, which accounted for 48.0% of the whole cohort. Most patients therefore did not change state within the observation period, largely because nearly half the cohort remained throughout in the state with the lowest support requirement. The 40.3% who did move are nonetheless a substantial subgroup, and it is precisely their course that a single phenotype per patient would have to average away.

These transitions were not one-directional. Patients moved from lower-support states into states requiring more organ support, and also back again; transitions between the two high-support states occurred in both directions. Notably, the most common destination on leaving neurological impairment- respiratory support was neurologically preserved-low support (9.2%), rather than the other high-support state. The course of these patients therefore does not follow a fixed severity ladder. (see Table 4)

**Table 4a.** Empirical state-transition probability matrix.

| From \ To | Neurological<br>impairment-renal<br>dysfunction | Neurological<br>impairment-respiratory support | Neurological<br>impairment-low<br>support | Neurologically<br>preserved-low<br>support |
| --- | --- | --- | --- | --- |
| Neurological<br>impairment-renal<br>dysfunction | 0.8526 | 0.0694 | 0.0126 | 0.0654 |
| Neurological<br>impairment-respiratory<br>support | 0.039 | 0.855 | 0.0143 | 0.0917 |
| Neurological<br>impairment-low<br>support | 0.0131 | 0.0255 | 0.7764 | 0.1851 |
| Neurologically<br>preserved-low support | 0.0119 | 0.0139 | 0.0197 | 0.9545 |

**Table 4b.** State episode duration (consecutive 6-hour windows)

| State | Median | Q25 | Q75 | N episodes |
| --- | --- | --- | --- | --- |
| Neurological impairment-renal dysfunction | 3.0 | 3.0 | 6.0 | 1556 |
| Neurological impairment-respiratory support | 3.0 | 1.0 | 7.0 | 2332 |
| Neurological impairment-low support | 3.0 | 1.0 | 4.0 | 1417 |
| Neurologically preserved-low support | 6.0 | 3.0 | 11.0 | 5998 |

The median duration of a single episode in neurologically preserved-low support was 36 h; for each of the other three states it was 18 h. The most common trajectories were as follows.

| **Trajectory** | **Patients** | **%** |
| --- | --- | --- |
| Neurologically preserved-low support throughout | 3058 | 48.0% |
| Neurological impairment-respiratory support throughout | 445 | 7.0% |
| Neurological impairment-respiratory support → neurologically preserved-low support | 371 | 5.8% |
| Neurological impairment-low support → neurologically preserved-low support | 188 | 3.0% |
| Neurological impairment-renal dysfunction throughout | 181 | 2.8% |

### The current state is associated with subsequent clinical events

After adjustment for age, sex, stroke subtype and the Charlson comorbidity index, the current state remained associated with organ-support escalation over the following 12 h and with ICU death within the first 72 h. Unadjusted and adjusted estimates are reported together in Table 5; adjustment moved the estimates only slightly and in no case reversed a direction.

**Table 5.**
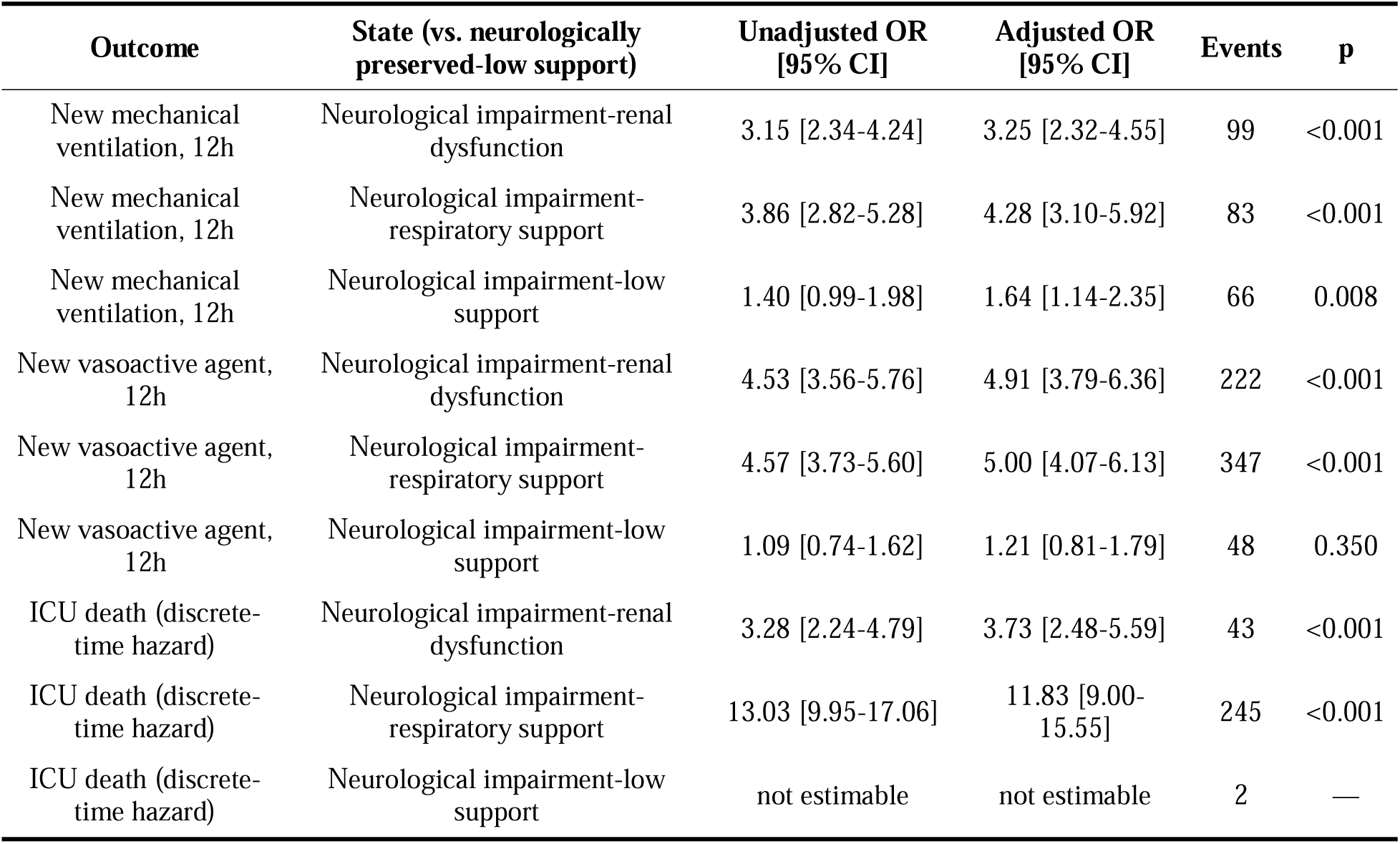
Adjusted association between current state and subsequent clinical events. Reference category: neurologically preserved-low support. Unadjusted estimates come from the same GEE specification with the covariates removed; adjusted estimates are additionally adjusted for age, sex, stroke subtype and Charlson comorbidity index. GEE, exchangeable working correlation, clustered by patient. An odds ratio is reported only where the state contributes at least 10 outcome events.

| Outcome | State (vs. neurologically preserved-low support) | Unadjusted OR [95% CI] | Adjusted OR [95% CI] | Events | p |
| --- | --- | --- | --- | --- | --- |
| New mechanical ventilation, 12h | Neurological impairment-renal dysfunction | 3.15 [2.34-4.24] | 3.25 [2.32-4.55] | 99 | <0.001 |
| New mechanical ventilation, 12h | Neurological impairment-respiratory support | 3.86 [2.82-5.28] | 4.28 [3.10-5.92] | 83 | <0.001 |
| New mechanical ventilation, 12h | Neurological impairment-low support | 1.40 [0.99-1.98] | 1.64 [1.14-2.35] | 66 | 0.008 |
| New vasoactive agent, 12h | Neurological impairment-renal dysfunction | 4.53 [3.56-5.76] | 4.91 [3.79-6.36] | 222 | <0.001 |
| New vasoactive agent, 12h | Neurological impairment-respiratory support | 4.57 [3.73-5.60] | 5.00 [4.07-6.13] | 347 | <0.001 |
| New vasoactive agent, 12h | Neurological impairment-low support | 1.09 [0.74-1.62] | 1.21 [0.81-1.79] | 48 | 0.350 |
| ICU death (discrete-time hazard) | Neurological impairment-renal dysfunction | 3.28 [2.24-4.79] | 3.73 [2.48-5.59] | 43 | <0.001 |
| ICU death (discrete-time hazard) | Neurological impairment-respiratory support | 13.03 [9.95-17.06] | 11.83 [9.00-15.55] | 245 | <0.001 |
| ICU death (discrete-time hazard) | Neurological impairment-low support | not estimable | not estimable | 2 | — |

Analyses of new organ support included only windows in which the patient was not already receiving that support. Because a state name summarizes dominant features rather than fixed criteria, 2324 windows (22.0%) assigned to neurological impairment-respiratory support were not receiving mechanical ventilation at that time point and were therefore eligible for the new-ventilation analysis. (see Table 5)

Death was counted only in the 6-hour window in which it occurred. Of the 714 ICU deaths in the cohort, 361 (51%) fell within the 72 h observation period; the remaining 353 patients were still in the ICU at 72 h and were censored in this analysis. What is estimated here is therefore the discrete-time hazard of death within each window of the first 72 h, not overall mortality across the whole ICU stay. Neurological impairment-low support contributed only 2 death events, and by the prespecified rule no odds ratio is reported for it.

At both the 24 h and the 48 h landmark, survival curves for patients in different states separated clearly (log-rank P<0.0001 at each landmark).

### The framework applies to all three stroke subtypes, but state distribution and outcomes differ

All four states occurred in all three subtypes, but in different proportions (χ²=1341.7, df=6, P<0.001). SAH had the most polarized distribution, with both the highest proportion of neurologically preserved- low support (68.0%) and the highest proportion of neurological impairment-respiratory support (21.9%); the distribution in AIS was comparatively even. Overall ICU mortality was 8.3% in AIS, 15.1% in ICH and 14.2% in SAH. (see Table 6a)

**Table 6a.** Window-level state distribution by stroke subtype (%)

| Subtype | neurologically preserved-low support | neurological impairment-low support | neurological impairment-renal dysfunction | neurological impairment-respiratory support |
| --- | --- | --- | --- | --- |
| AIS | 63.9 | 9.1 | 13.7 | 13.2 |
| ICH | 59.9 | 7.6 | 10.9 | 21.7 |
| SAH | 68.0 | 3.4 | 6.7 | 21.9 |

**Table 6b.**
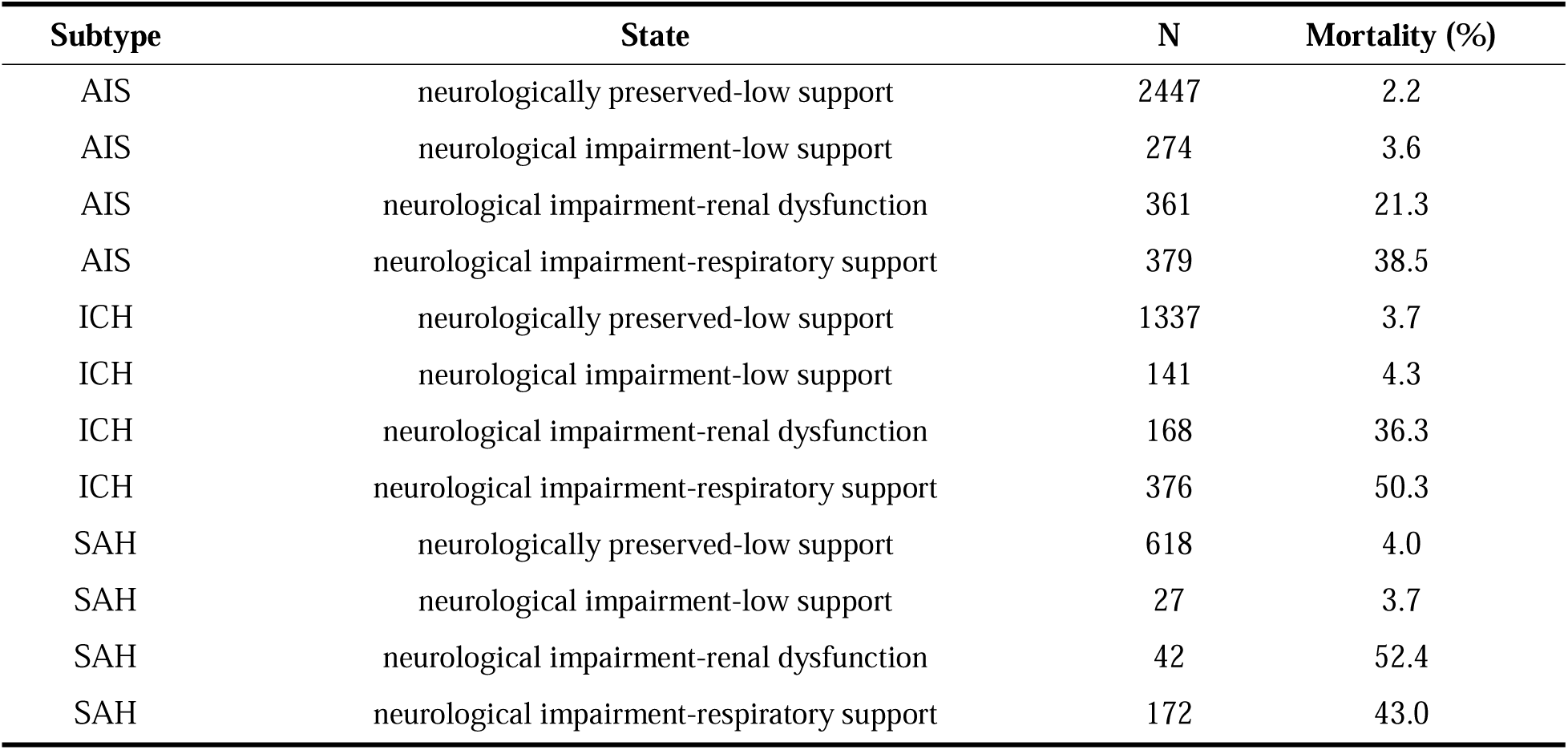
ICU mortality (%) by subtype and last observed state (n)

A further descriptive analysis showed that ICU mortality was not the same across subtypes even for patients ending in the same state. Among patients whose last observed state was neurological impairment- renal dysfunction, ICU mortality was 21.3% in AIS, 36.3% in ICH and 52.4% in SAH. The corresponding figures for neurologically preserved-low support were 2.2%, 3.7% and 4.0%, and for neurological impairment-low support 3.6%, 4.3% and 3.7%. This pattern suggests that the clinical meaning of a state may be influenced by the underlying stroke mechanism, but confirming that would require a formal interaction analysis and external validation.

### Robustness of the state structure

#### Reproducibility in the held-out test set

When the model fitted in the training set was frozen and applied directly to the held-out test set, the distribution of the four states was almost unchanged: 63.0% versus 64.1%, 7.9% versus 7.4%, 11.9% versus 11.4%, and 17.2% versus 17.1%, a maximum absolute difference of 1.1 percentage points.

#### Reproducibility across admission eras

MIMIC-IV spans 2008–2022, and practice changed over that period: ICU mortality in this cohort fell from 13.8% in patients admitted in 2008–2010 to 7.6% in 2020–2022. Admission era is therefore not a null contrast, and it is the closest approximation to external validation available within a single database.

Applying the frozen primary model to each era, the mix of states shifted with that secular trend. The share of windows in neurologically preserved-low support rose from 57.7% in 2008–2010 to 69.4% in 2020–2022, while neurological impairment-respiratory support fell from 19.3% to 11.4% and neurological impairment-renal dysfunction from 15.6% to 8.9%.

The behavior of the states themselves was far more stable than their prevalence. Self-transition probability across the five eras varied between 93.5% and 96.8% for neurologically preserved-low support, 73.5% and 81.4% for neurological impairment-low support, 83.8% and 86.7% for neurological impairment-renal dysfunction, and 82.1% and 87.6% for neurological impairment-respiratory support.

Refitting the model from scratch within an early era (2008–2013, 2500 patients) and a late era (2017–2022, 2550 patients) recovered a four-state structure in both, with restart stability of 0.936 and 0.959 respectively. Agreement with the primary assignment was ARI 0.860 in the early era and 0.899 in the late era.

Across admission eras, state prevalence changed substantially, whereas state profiles and transition persistence were comparatively stable.

#### Prespecified sensitivity analyses

Of the ten prespecified sensitivity analyses, nine yielded a stable four-state solution, with agreement against the primary model ranging from ARI 0.629 to 0.965. Among the four analyses specified during execution, the three alternative treatments of the GCS verbal score reproduced the structure (ARI 0.761– 0.831). Robust standardization did not produce a stable multi-state solution at any setting tested. Agreement was not uniform across the ten, and the analysis with the weakest agreement is informative rather than merely weak: it is described below. (see Table 7)

**Table 7.** Sensitivity analysis summary.

| **Analysis** | **Specified** | **ARI vs.**<br>**primary model** | **Interpretation** |
| --- | --- | --- | --- |
| Exclude patients sedated in >50% of their windows | Prespecified | 0.965 | The states are not principally an artefact of who was sedated |
| Add panel-level missingness indicators | Prespecified | 0.955 | State assignment does not depend on lab-ordering behaviour |
| ICU stay $\geq 24$ h (original protocol threshold) | Prespecified | 0.946 | n=5,726; relaxing the threshold did not change the structure |
| Exclude all four treatment variables (pre-specified most important) | Prespecified | 0.935 | States are driven by physiology, not treatment decisions |
| AIS patients only | Prespecified | 0.895 | Not an artifact of pooling three subtypes |
| Within-window mean instead of last value | Prespecified | 0.875 | Aggregation rule is not decisive |
| 12-hour time windows | Prespecified | 0.824 | Structure is not an artifact of window length |
| Complete cases only: all 17 continuous variables measured | Prespecified | 0.772 | Retained windows are neither contiguous nor representative; a lower bound |
| Exclude all eight laboratory variables | Prespecified | 0.754 | The renal dysfunction state dissolves; it is defined by creatinine and BUN |
| No forward fill, laboratory variables removed | Prespecified | 0.629 | Not estimable with the full feature set (no restart converged in 50) |
| GCS verbal imputed to the training median (5) | Added during execution | 0.831 | The earlier specification; reproduces the earlier state profile |
| GCS verbal regression-imputed from eye + motor | Added during execution | 0.828 | Steepest mortality gradient of the three verbal handlings |
| GCS verbal floor-coded to 1 | Added during execution | 0.761 | Compresses the mortality gradient; worst of the three |
| Robust (median/IQR) scaling | Added during execution | not estimable | EM collapsed to a single state at every setting tested |

With all treatment variables removed, state assignment remained highly concordant with the primary model (ARI 0.935), which argues against the states being driven mainly by treatment. Adding laboratory missingness indicators gave an ARI of 0.955, arguing likewise against the structure being driven mainly by testing patterns.

The analysis with the weakest agreement was the removal of all laboratory variables (ARI 0.754). This model still returned four states, but they were not the same four: neurological impairment-renal dysfunction no longer appeared. Rather than undermining the structure, this identifies what defines that state — renal measurements, which vital signs cannot substitute for. All three alternative treatments of the GCS verbal score reproduced the four-state structure (ARI 0.761–0.831).

Two further analyses named in the design document bear on specific concerns about the state definition. Because GCS-EM carries the neurological dimension and sedation depresses it independently of brain injury, the model was refitted after excluding the 931 patients (14.6%) who received a continuous sedative infusion in more than half of their observed windows. Sedation depth was proxied by infusion status, RASS not having been extracted. Agreement with the primary model was ARI 0.965, and the four states retained their profiles and their mortality gradient (2.4% to 54.2%); the states are therefore not principally an artefact of who was sedated.

A complete-case analysis used only the 8048 training windows (18.6%) in which all 17 continuous variables were actually measured, with no forward filling and no median imputation anywhere. Four states were again recovered, with high restart stability (0.996) and agreement with the primary model of ARI 0.772. Two features of this comparison should be read together with that figure. Complete windows are not contiguous within a patient, so the sequences contain gaps that the model treats as adjacent; state assignment is therefore comparable with the primary model but transition quantities are not, and none are reported here. And complete windows are not a random subset: a window is complete when a full laboratory panel was drawn, which happens more often in sicker patients, so this subset is more heavily ventilated and sedated than the cohort as a whole. The agreement of 0.772 is thus a lower bound obtained on a deliberately unfavourable subsample.

A further analysis, named in the design document, removed forward filling entirely, imputing every unmeasured window with the training-set median. With the full feature set this specification is not estimable under the present emission model: without forward filling each of the eight laboratory variables takes a single value in 66–71% of windows, and a diagonal-covariance normal emission cannot be fitted to a variable that is mostly a point mass. No restart converged in 50 attempts, at any variance floor between 0.05 and 0.5. Removing the laboratory variables removes those point masses, and the specification then converges with a restart stability of 0.899; agreement with the primary model was ARI 0.629, and the four recovered states retained a mortality gradient from 2.8% to 33.7%. The question this analysis was designed to address — whether the states are an artefact of imputation — is answered more directly by the analysis that removes the laboratory variables outright while retaining forward filling (ARI 0.754), reported above. Refitting every model with uncapped age, which affects the 73 patients whose derived age exceeds 91 years, changed the state odds ratios by at most 0.008 in absolute terms.

## 4. Discussion

The most important finding of this study is not that patients with acute stroke in the ICU can be sorted into four groups, but that they move between four clinically interpretable states. Observed at 6- hour intervals, 40.3% of patients changed state at least once during the first 72 h, and the state a patient currently occupied was associated with organ-support escalation over the following 12 h and with death within 72 h. Transitions did not run in one direction: the most common destination on leaving neurological impairment-respiratory support was neurologically preserved-low support, not the other high-support state. The four states are therefore not a graded scale from mild to severe, but distinct clinical situations that a patient may enter and leave during the ICU course.

This is what separates the present work from trajectory phenotyping, which has been applied to critically ill patients with ischemic stroke to identify subgroups with distinct longitudinal vital sign patterns and treatment response[3], and more recently to the Glasgow Coma Scale across three ICU databases, where patient-level trajectory classes added prognostic information beyond baseline variables[4]. That work and this one differ in what they set out to do. Their object was to improve outcome prediction, and they quantified the gain. Ours was to establish, before any prediction is attempted, a multidimensional space in which the same patient can occupy different clinically distinct states at different times; the incremental predictive value of state sequences is a separate question and is the intended next study. The two also differ in what is being represented: a single physiological score summarized into one class per patient, against twenty-one neurological, physiological and organ-support variables resolved into a state per 6-hour window. Such studies use longitudinal data but resolve to one label per patient, assigned from the pattern over an interval. The representation here is different in kind: the patient occupies a state at each 6-hour window, and the object of analysis is the movement between those states. The distinction is not merely technical. In this cohort a fixed phenotype would have had to absorb the 40.3% of patients who changed state, and would have concealed exactly the events that matter at the bedside: weaning, and overnight deterioration. A risk model answers how likely an outcome is; a phenotype answers what kind of patient this is; a state-transition model answers where the patient is now and which way the course is moving. These are complementary rather than competing.

The four states also show that neurological impairment is not a single pattern. GCS-EM was reduced in all three impaired states, but the systemic picture differed: some patients had reduced responsiveness yet rarely needed organ support, some showed marked renal abnormality alongside it, and some were dominated by ventilation and sedation requirements. A similar level of neurological responsiveness can therefore occur against quite different systemic backgrounds, and those differences were related to subsequent organ-support escalation and death. That the renal dysfunction state disappeared when laboratory variables were removed shows what defines it: renal measurements contribute information that vital signs cannot supply. Understood this way, the states form an interpretable layer between raw monitoring data and clinical outcome — they condense scattered neurological, physiological and treatment information into a stage of illness that can be reasoned about clinically, while preserving how a patient changes over time.

The structure was stable across several prespecified checks. State prevalence was almost unchanged when the frozen training model was applied to the held-out test set (maximum difference 1.1 percentage points); removing all four organ-support variables left assignment highly concordant with the primary model (ARI 0.935), which matters because treatment is part of the state definition; and adding laboratory missingness indicators changed little (ARI 0.955), arguing against the structure reflecting testing behavior. The most demanding of these checks was across admission eras. ICU mortality in this cohort fell from 13.8% to 7.6% between 2008–2010 and 2020–2022, and the mix of states moved with it — the preserved-low support state rose from 57.7% to 69.4% of windows. Yet refitting the model separately in an early and a late era recovered the four states in both, with agreement of ARI 0.860 and 0.899 against the primary assignment, and self-transition probabilities varied by only a few percentage points across the whole period. What changed over fifteen years was how many patients occupied each state, not what the states are. Two constraints on interpretation should be stated plainly. What was observed is a prognostic association between state and subsequent events, not a causal relationship, and nothing here supports an inference that a given treatment moves a patient from one state to another. Separately, the same framework described all three stroke subtypes, but in an exploratory analysis mortality for the same last observed state was not consistent across them; a state is therefore not a substitute for the underlying stroke mechanism, and establishing that difference would require a formal interaction analysis.

Several limitations remain. The data come from a single academic medical center. Reproducibility across admission eras is reassuring but is not external validation: practice at one institution over time is not the same test as practice at another institution, and state prevalence in particular may reflect local admission and treatment thresholds. MIMIC-IV lacks the NIHSS and systematic neuroimaging severity measures, so the neurological dimension reflects responsiveness as observable in the ICU. GCS-EM can also be depressed by sedation rather than by brain injury, although excluding the most heavily sedated patients left the structure essentially unchanged (ARI 0.965). Each 6-hour window is represented by summary values, which cannot preserve short-term fluctuation within the window. Several laboratory variables had high window-level missingness; the missingness-indicator analysis gave no sign that this dominated state assignment, but this needs confirmation elsewhere. Patients with an ICU stay shorter than 12 h were excluded, so the earliest deaths may be under-represented. Two further constraints follow from the model specification itself. The emission model treats all continuous features as normally distributed, including the GCS eye and motor scores, which are ordinal with only four and six levels; this is a pragmatic choice that permits joint modelling with the physiological variables but does not respect the measurement scale of those variables, and it is the proximate cause of two failures reported here — the collapse of robust standardization and the non-estimability of the no-forward-fill specification. A mixed- emission model with ordinal components for the neurological subscores is the more principled specification and is left to future work. Relatedly, laboratory values are measured in only about a third of windows, so the renal dysfunction state is defined by variables that are imputed in most of the windows to which it is assigned. A complete-case analysis using only fully measured windows recovered four states with moderate agreement (ARI 0.772), but that subset is neither contiguous in time nor representative of the cohort, so the dependence of the recovered structure on how sparsely measured laboratory variables are represented over time is reduced rather than removed. The four states are not equally well supported by these data. Neurological impairment-low support is the smallest at 7.8% of windows, contributed only 2 discrete-time death events so that no odds ratio could be estimated for it, and was the least well recovered of the four when the organ-support variables were removed (74.6% of its windows retained, against 86.5-99.7% for the other three). Whether all four states are recoverable outside this cohort, or whether the three defined by a positive feature — preserved responsiveness, renal dysfunction, respiratory support — constitute the more robust structure, is a question that external validation must answer rather than confirm. Finally, this study does not yet predict which state a patient will enter next; establishing a stable, interpretable state space and describing movement within it was the prerequisite. The next steps are to use prior state sequences to predict transitions over the following 6–12 h, and to test in external data whether these four states can be identified again and whether their clinical profiles and transition patterns hold.

## Conclusions

Using longitudinal data from the first 72 h of ICU care in 6368 patients with acute stroke, we identified four reproducible and clinically distinguishable dynamic states. About 40% of patients changed state during the observation period. For that substantial subgroup, the early ICU course is better represented as a sequence of changing states than as a single fixed phenotype. After adjustment for baseline characteristics, the current state remained associated with subsequent organ-support escalation and with ICU death within the first 72 h. The next steps are external validation and prediction of the transitions a patient may undergo over the following 6–12 h.

## Ethics statement

This study is a retrospective analysis of MIMIC-IV v3.1, a de-identified database distributed through PhysioNet under credentialed access. Collection of the underlying patient data and creation of the research resource were reviewed by the Institutional Review Board of Beth Israel Deaconess Medical Center, which granted a waiver of informed consent and approved the data sharing initiative[4]. The corresponding author completed the CITI Program course Data or Specimens Only Research and signed the PhysioNet Credentialed Health Data Use Agreement (v1.5.0) before accessing the data. As this work involved only secondary analysis of an existing de-identified public dataset, no additional institutional review or informed consent was required. The study was conducted in accordance with the principles of the Declaration of Helsinki.

## Consent for publication

Not applicable.

## Availability of data and materials

The dataset analyzed in this study is publicly available through PhysioNet (https://physionet.org/content/mimiciv/3.1/). Analysis code will be made available upon reasonable request.

## Author contributions

P.L.: Conceptualization, Data curation, Formal analysis, Methodology, Software, Validation, Visualization, Writing – original draft, Writing – review and editing. Y.X.: Conceptualization, Methodology, Formal analysis, Writing – review and editing. Y.Z.: Investigation, Validation, Writing – review and editing; adjudicated the clinical characterization of the identified states while blinded to outcome data. All authors read and approved the final manuscript.

## Declaration of generative AI and AI-assisted technologies

During the preparation of this work the authors used generative AI tools (OpenAI ChatGPT; Anthropic Claude / Claude Code) to assist with implementing the analysis code, generating figures, and drafting manuscript text. All study design decisions, variable and cohort definitions, analytic choices, and interpretations of the results were made by the authors. The corresponding author independently re-derived and verified every numerical result reported in this manuscript against the underlying data and analysis outputs, and the clinical co-author independently adjudicated the clinical characterization of the identified states. The authors reviewed and edited all AI-assisted output and take full responsibility for the content of the publication. No AI tool is listed as an author.

## Supporting information

Statistical Analysis Plan

## Data Availability

The dataset analyzed in this study is publicly available through PhysioNet (https://physionet.org/content/mimiciv/3.1/) to credentialed users who have completed the required training and signed the data use agreement. The data cannot be redistributed by the authors. Analysis code will be made available upon reasonable request.

## Acknowledgments

The authors thank the School of Public Health, Fudan University, and the Institute of Global Health, University of Geneva, for institutional support.

---

**Fig. 1.**
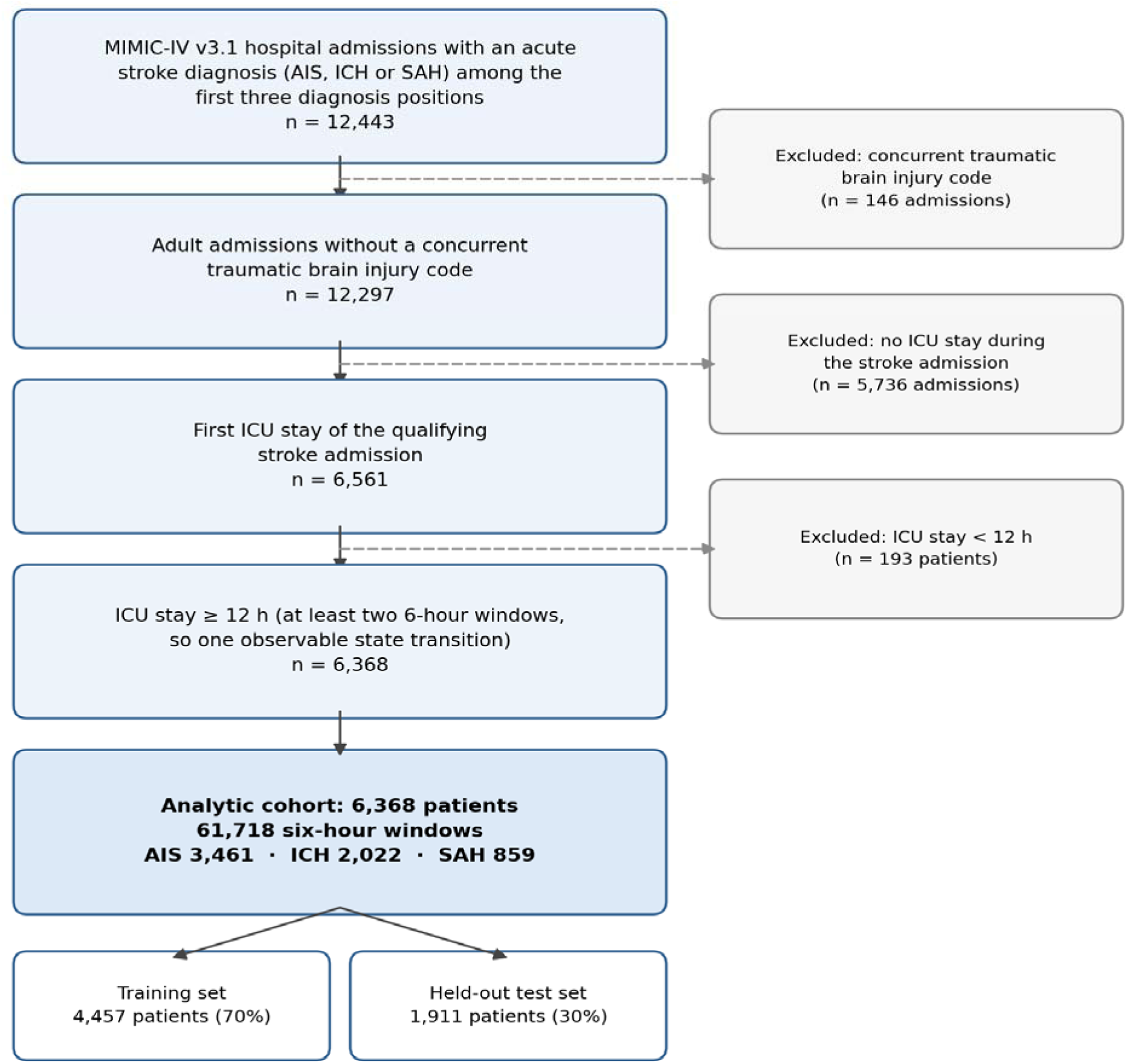
Study population flowchart. Patient selection from MIMIC-IV v3.1, showing inclusion and exclusion criteria and the final analytic cohort.

**Fig. 2.**
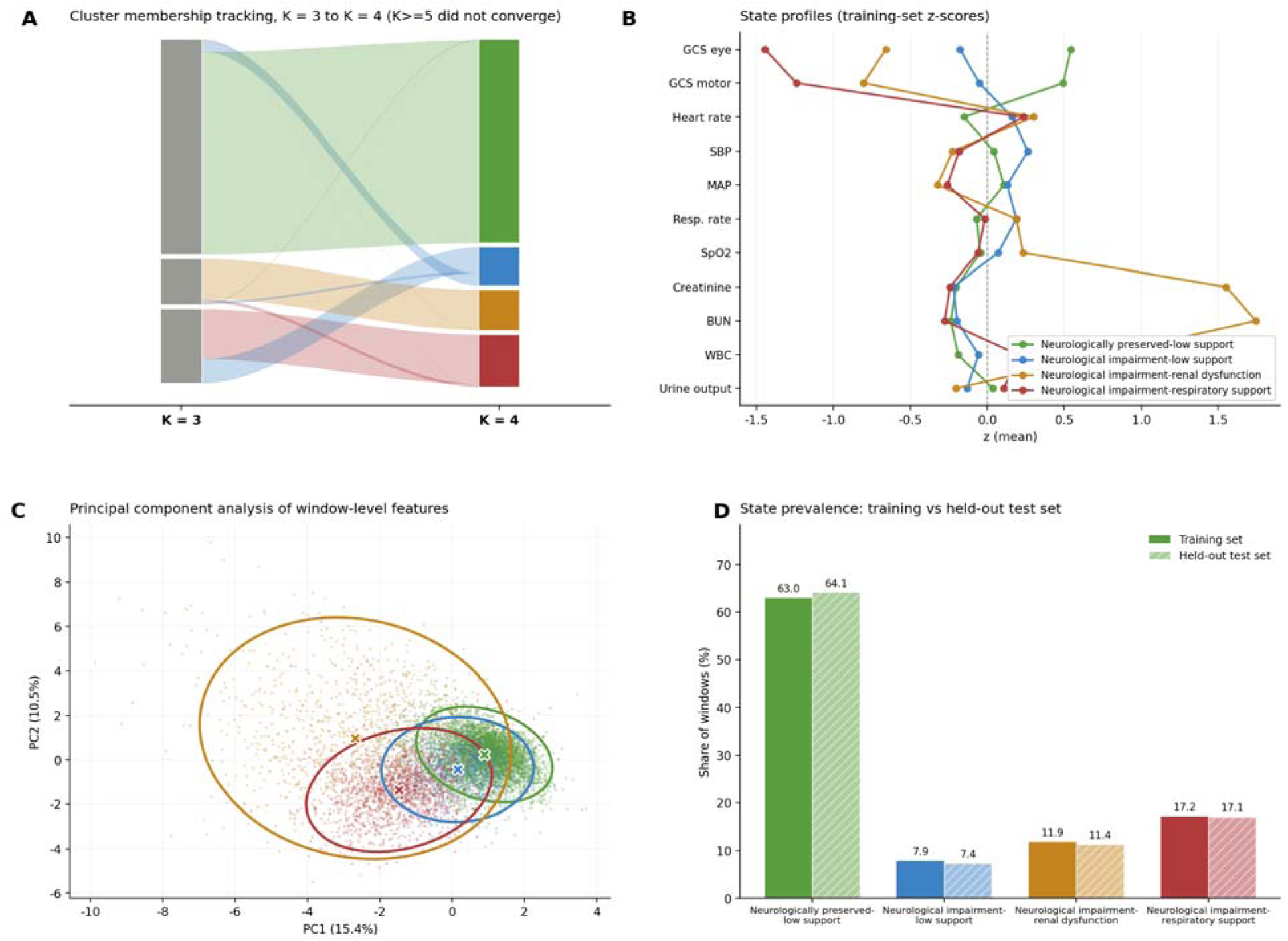
Identification and characterization of the dynamic clinical states. (A) Cluster membership tracking across candidate state counts (K = 3 to 5); the K = 4 solution is colored, adjacent solutions in grey. (B) State profiles, expressed as training-set z-scores of key variables. (C) Principal component analysis of the 21 window-level features, colored by assigned state, with 95% confidence ellipses and state centroids. (D) State prevalence in the training set and in the held-out test set, by state; the frozen training model is applied unchanged to the test set.

**Fig. 3.**
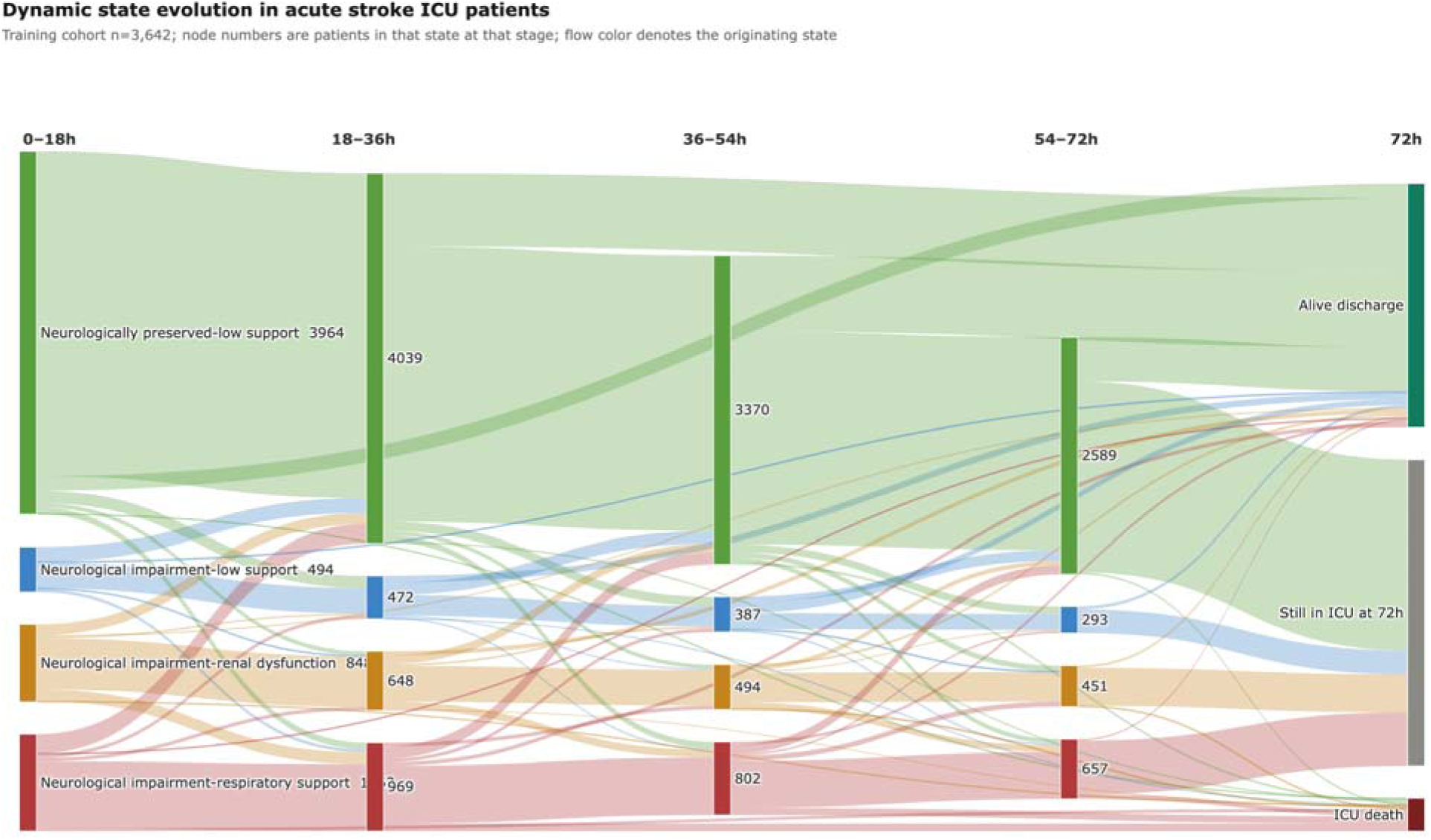
Dynamic state evolution pathways. Sankey diagram of state transitions across four consecutive 18-hour stages in the full cohort (n=6368). Node height is proportional to the number of patients in that state; flow color denotes the originating state. Terminal nodes represent ICU death, live ICU discharge, and continued ICU stay at the 72-hour cutoff.

**Fig. 4.**
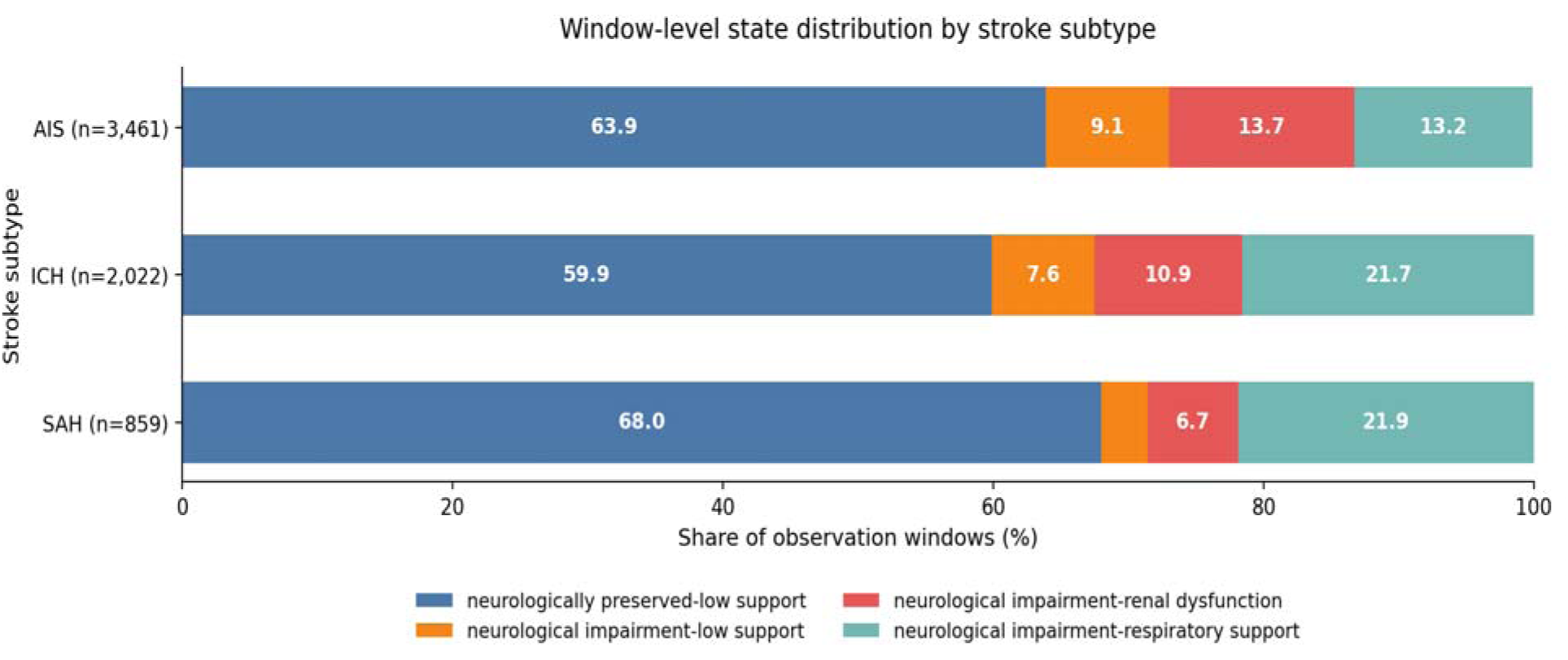
Window-level state distribution by stroke subtype. AIS, ischemic stroke; ICH, intracerebral hemorrhage; SAH, subarachnoid hemorrhage.

**Fig. 5.**
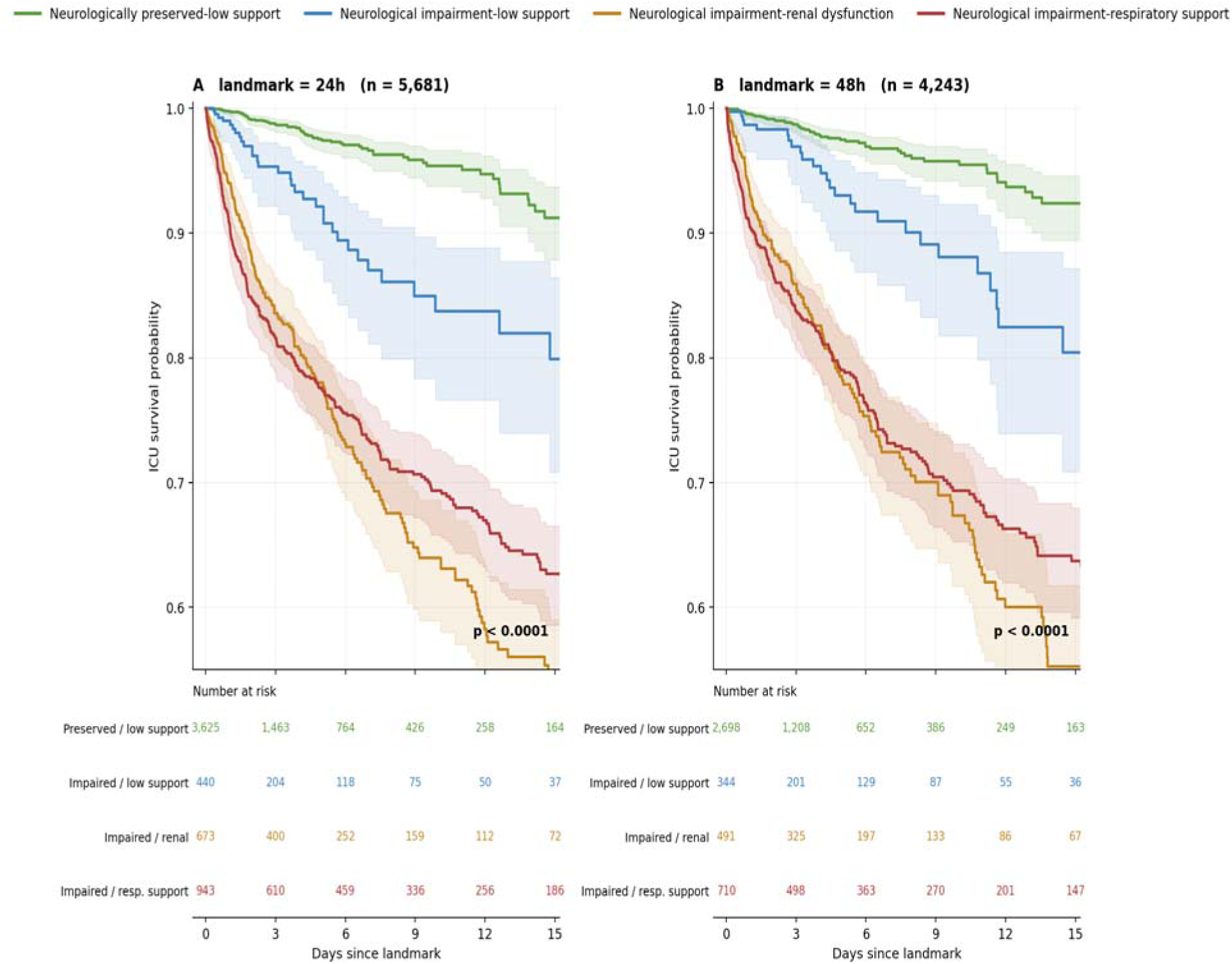
Landmark Kaplan-Meier analysis of ICU survival by dynamic state. Because state occupancy is time-varying, patients still in the ICU at (A) 24 hours and (B) 48 hours were stratified by the state occupied in that window and followed from that landmark onward, reducing immortal-time bias. Shaded bands are 95% confidence intervals; p values are from the log-rank test.

**Supplementary Fig. S1.**
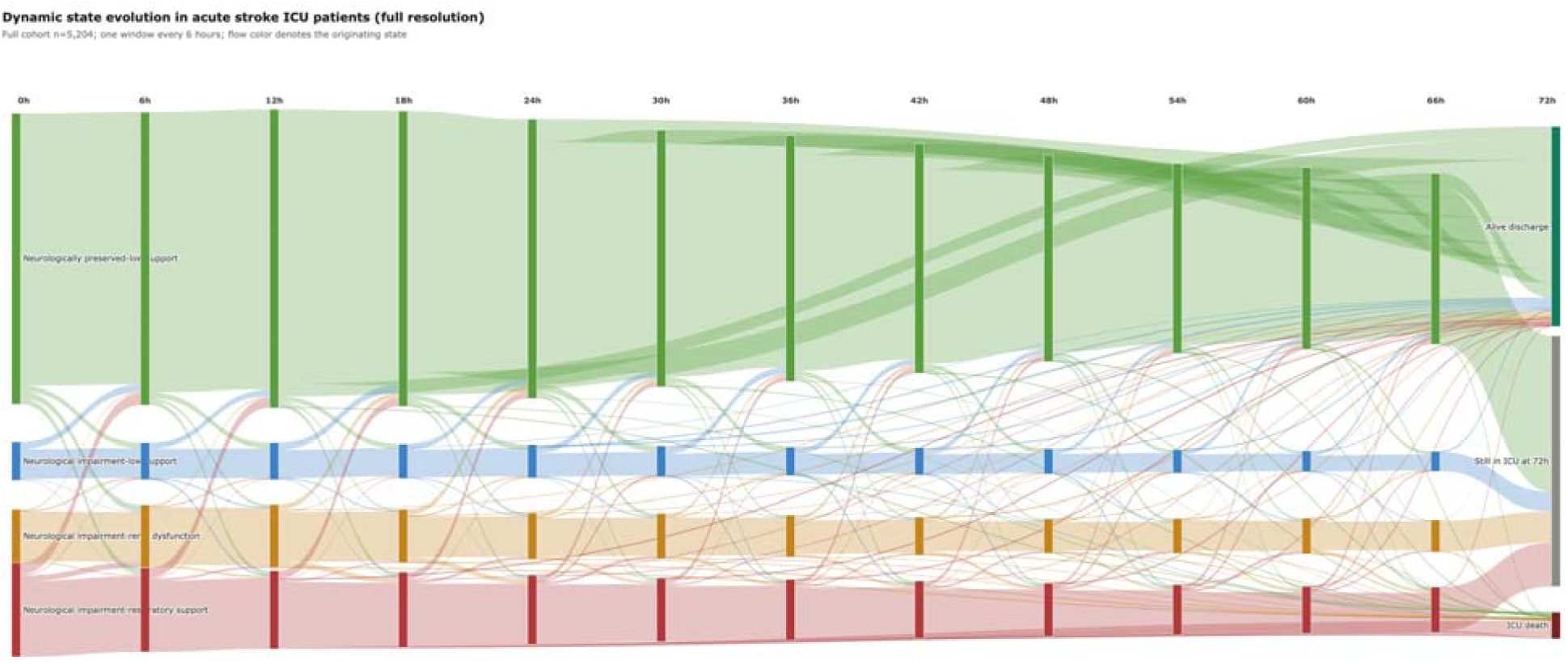
Full-resolution state evolution pathways across all twelve 6-hour windows in the full cohort (n=6368). Vertical ordering of states is fixed across columns to make transitions readable.

**Supplementary Fig. S2.**
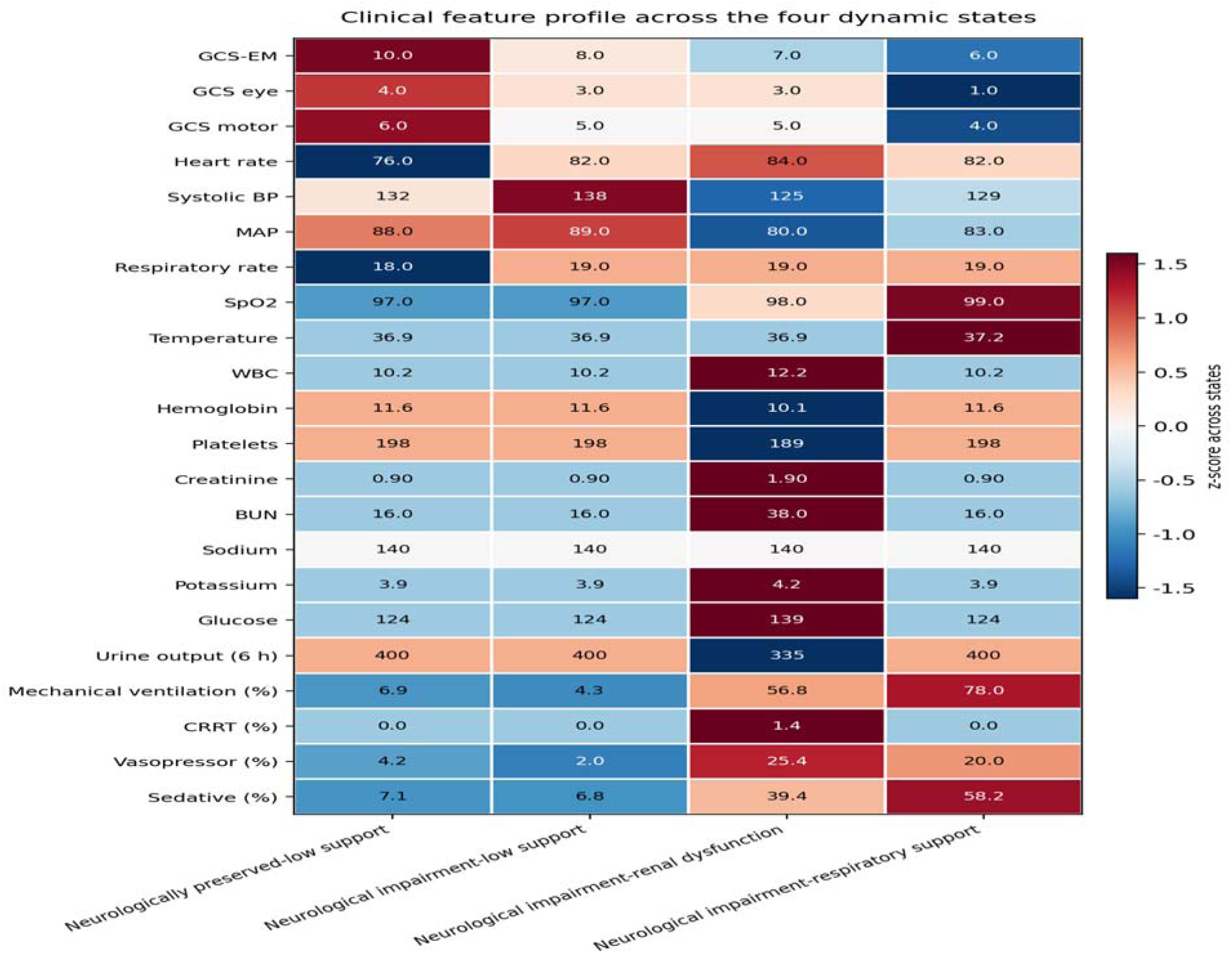
Clinical feature heatmap across the four dynamic states. Cell values are raw medians; color encodes the z-score of each variable across the four states.

**Supplementary Fig. S3.**
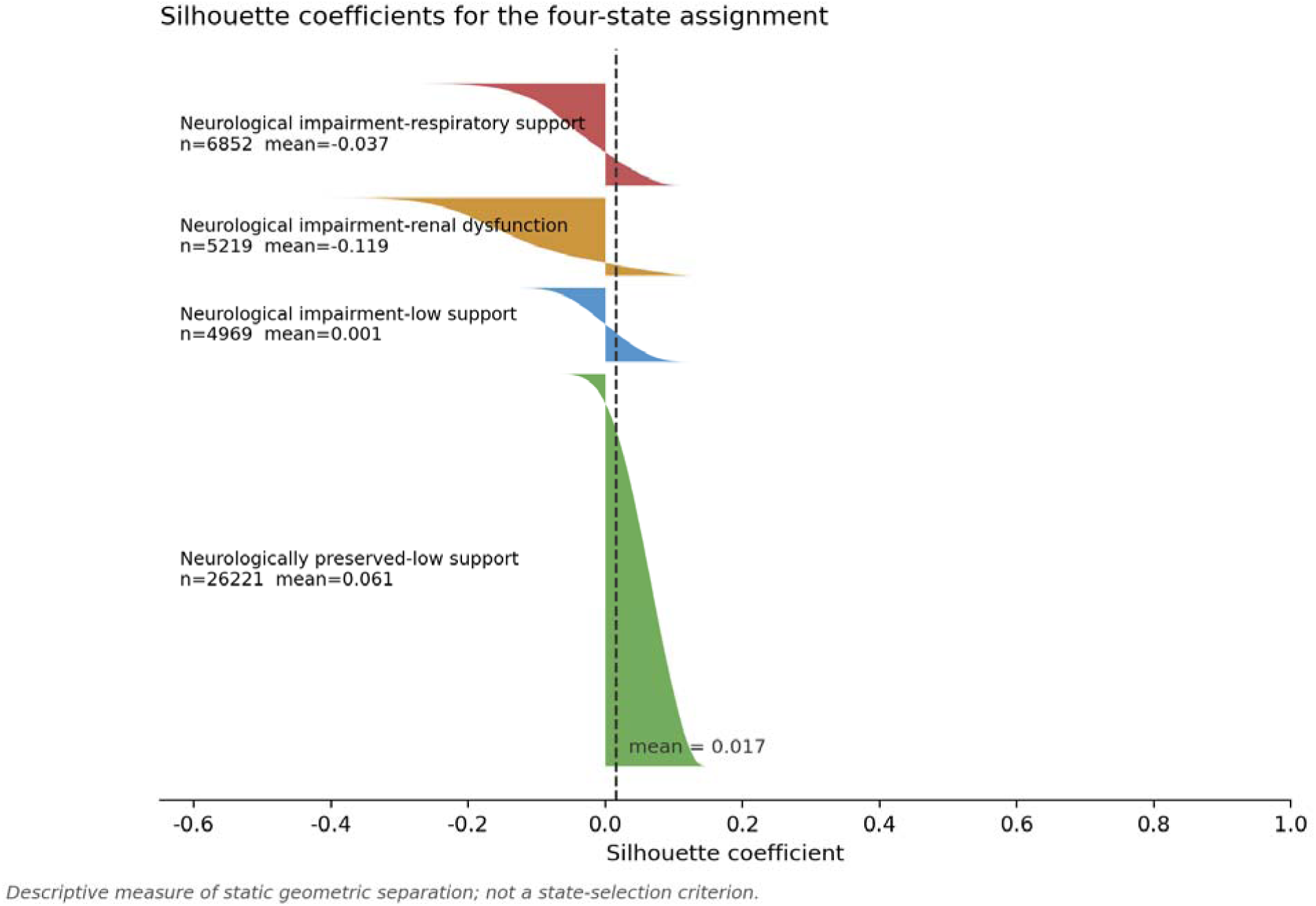
Silhouette coefficients for the four-state assignment, computed over the full training set. Silhouette is reported as a descriptive measure of static geometric separation in the raw feature space. It was not a state-selection criterion: it evaluates each window as an independent point and is therefore blind to the transition structure that distinguishes these states from clusters.

## STROBE Statement — checklist of items for cohort studies

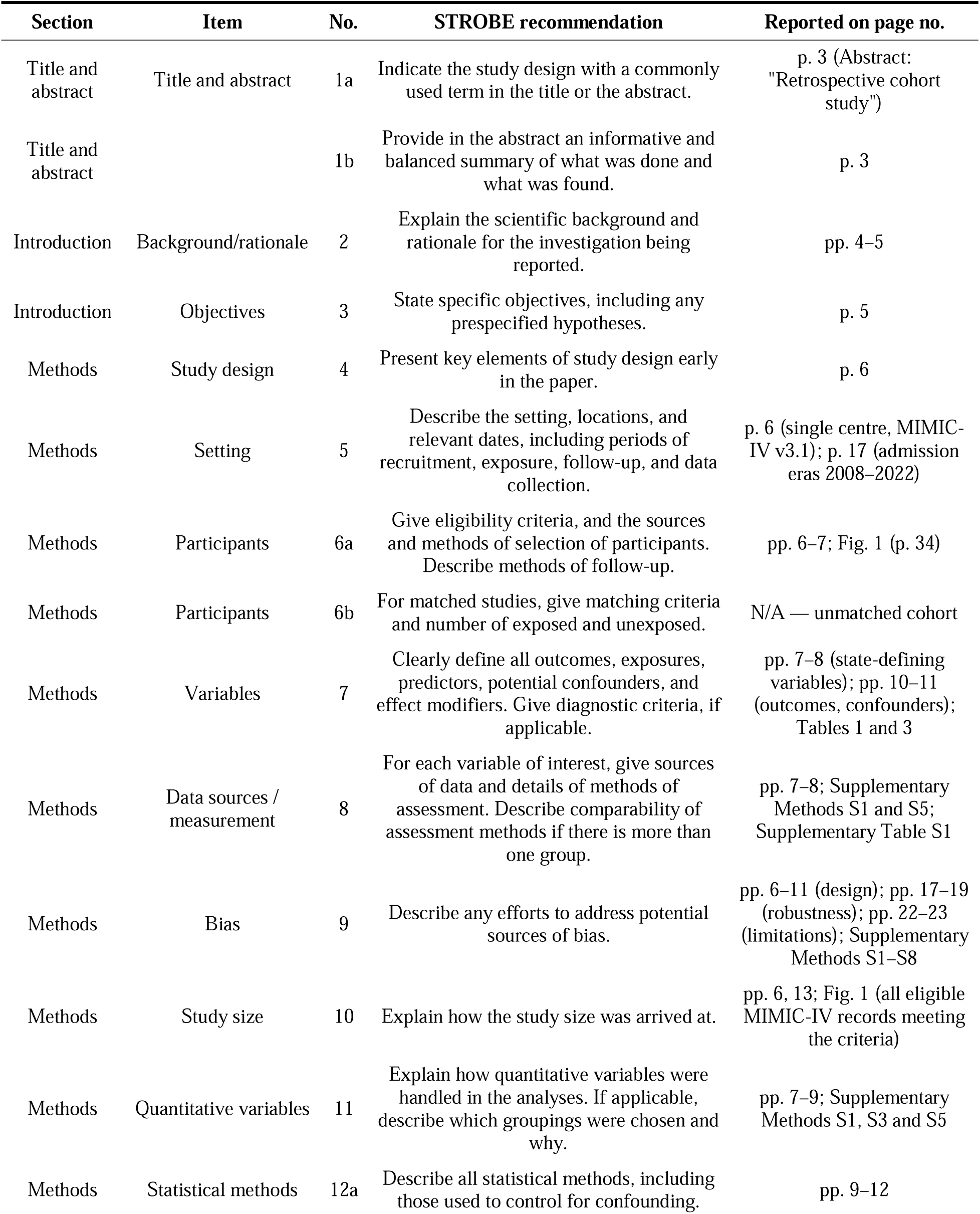
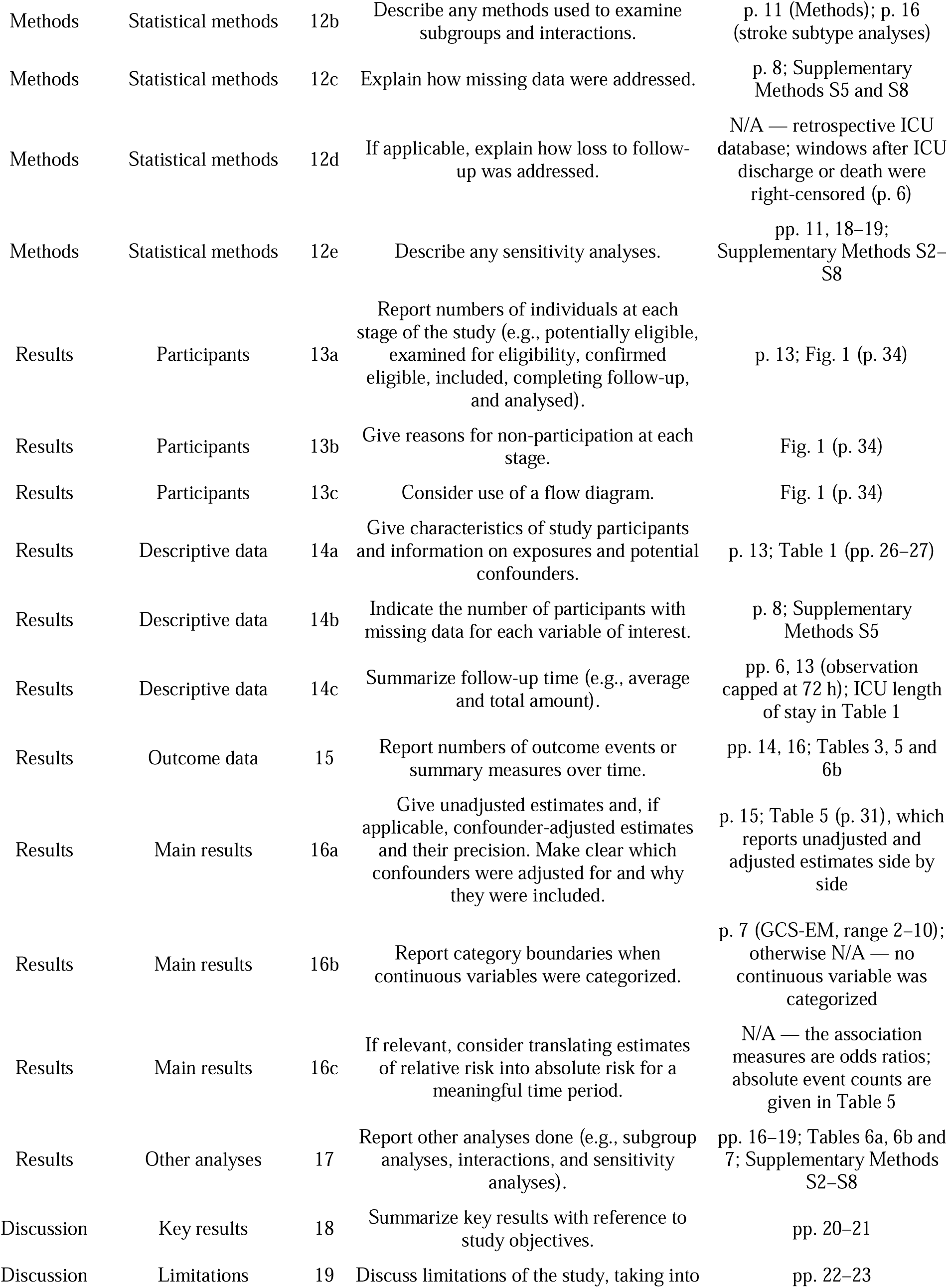
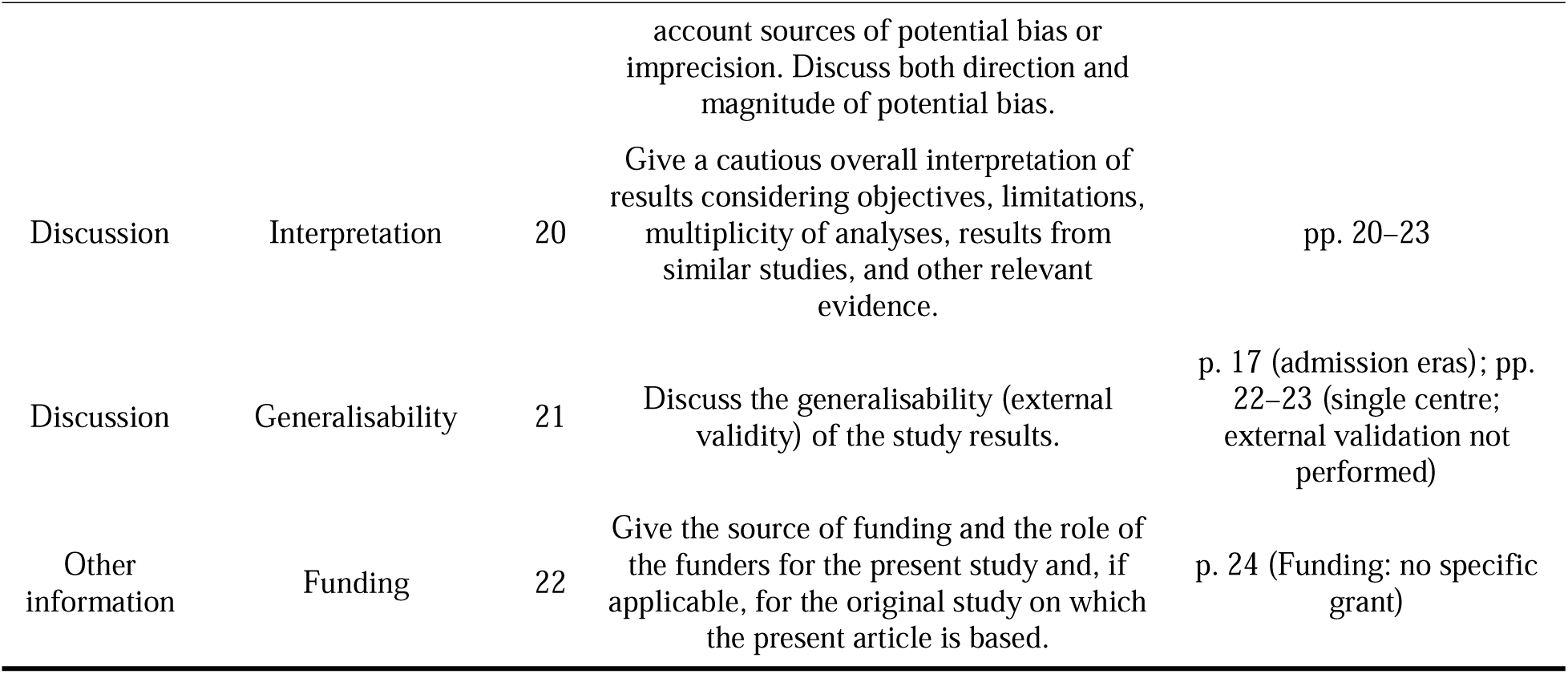

