## Supplementary Material for "Dynamic Clinical States and Transitions During the First 72 Hours of Intensive Care After Acute Stroke"

*Supplementary Methods S1 · Two-tier data quality control*

Two tiers of quality control were applied to the extracted tables. The first sets physiologically impossible values to missing. The second flags values that are extreme but physiologically possible; these are retained and listed in the quality-control report. Across 18 continuous variables, 372 values were removed under the first rule — a heart rate of 790 beats/min, a mean arterial pressure of 75149 mmHg, an oxygen saturation of 3333330%, and similar entry or sensor artefacts.

Urine output shows why the two tiers must be distinguished. An earlier version of the pipeline deleted any 6-hour window totalling more than 5000 mL. Inspection of the source records showed three things. First, there was no instance of a cumulative value being recorded as an interval value: the 99.99th percentile of a single record was 2229 mL and the maximum 4385 mL, both plausible for one void or drainage. Second, only 15 windows exceeded 5000 mL, 0.030% of all windows. Third, those windows clustered in consecutive windows of a small number of patients, consistent with sustained polyuria — commonly from osmotic diuresis after mannitol or hypertonic saline given for intracranial pressure, or from central diabetes insipidus after severe brain injury. Both have clear clinical meaning in a stroke ICU population. On that basis 5000 mL is now a manual review threshold, and the deletion boundary is set at a physiologically impossible level.

*Supplementary Methods S2 · Choice of the number of states*

***S2.1 What was fixed in advance***

The dimensions on which candidate models would be judged were written into the study design document before any model was fitted, and are reproduced in the accompanying Prespecified Modeling Strategy. They were: statistical fit; the share of patients and windows in each state; whether any state was very small; whether states differed clinically in a substantive way; whether results were stable across random initializations; and whether clinicians could interpret them.

The design document also stated, in advance, that the number of states must not be chosen on the Bayesian information criterion alone. The reason given was that in latent class trajectory analysis small changes in model specification can materially alter both the recovered classes and their apparent clinical associations, so stability and clinical interpretation must enter the decision. It stated explicitly that the final number of states is not determined by statistical indices alone.

This is a set of evaluation dimensions with one stated exclusion. It is not a deterministic decision rule: no weighting between criteria was fixed, and the final choice was a judgment across all six. We state this distinction rather than presenting the selection as mechanical.

***S2.2 Candidate models***

| **States** | **Log-likelihood** | **BIC** | **Restart stability (ARI)** | **Restarts converged** | **Minimum state occupancy** |
| --- | --- | --- | --- | --- | --- |
| 3 | −721,152 | 1,443,606 | 0.983 | 7/10 | 13.5% |
| 4 | −717,281 | 1,436,345 | 0.995 | 7/10 | 7.9% |
| 5 | −676,187 | 1,354,657 | not estimable | 1/10 | 9.1% |
| 6 | — | — | not estimable | 0/10 | — |

***S2.3 Criterion by criterion***

**Statistical fit.** On BIC alone the five-state model is clearly preferred, and would have been selected. This is stated plainly because it is the single strongest argument against the chosen model.

**Convergence and restart stability.** The five-state model converged in 1 of 10 restarts. Increasing to 50 restarts produced one further convergent fit and no more; the six-state model likewise converged once in 50. Restart stability is defined as the agreement between the best and the second-best solution, so with only one solution the quantity does not exist — it is not low, it is undefined. The five-state model therefore fails this criterion outright rather than scoring poorly on it. Among the models that did converge repeatedly, the four-state model was the most stable (ARI 0.995 versus 0.983 at three states).

**State occupancy.** The minimum state occupancy was 7.9% at four states, well above the 1–2% level that the plan set as a trigger to reconsider. No candidate produced a degenerate state.

**Clinical distinctness.** The additional state in the five-state solution comes from subdividing the largest state. The two resulting substates have the same median GCS-EM (10), the same median creatinine (0.90 mg/dL) and the same median urine output; ICU mortality is 2.8% and 3.1%. They differ materially only in the proportion of windows receiving mechanical ventilation (10.2% versus 5.7%). The BIC advantage therefore comes from resolving the largest state more finely, not from identifying a new clinical phenotype. This fails the prespecified requirement that states differ clinically in a substantive way.

**Interpretability.** The four states were adjudicated by a critical care nurse scientist blinded to outcome data (Supplementary Methods S4); the five-state solution offered no additional state that clinicians could describe distinctly from its parent.

***S2.4 Summary***

The four-state model was selected because it was the only candidate to satisfy all six criteria. The five-state model had the better statistical fit and failed on stability and clinical distinctness; the three-state model satisfied every criterion but was less stable and merged states that the four-state model separates on renal function.

*Supplementary Methods S3 · Choice of standardization*

Robust standardization using medians and interquartile ranges was prespecified, on the grounds that physiological variables in critically ill patients are strongly right-skewed: in the training set the skewness of creatinine, blood urea nitrogen and glucose was 6.87, 3.75 and 3.31 respectively.

It could not be used with this emission model. At every candidate state count from 3 to 6, at every random restart, and at every variance floor tested between 0.01 and 0.2, expectation-maximization degenerated to a single-state solution in which all training windows were assigned to one state. The likely mechanism is that after interquartile-range scaling the GCS eye and motor scores become discrete variables taking only 4 and 6 distinct integer-spaced values, while still being modeled as continuous normal emissions, which drives the per-state variance on those features toward collapse. An ordinal emission distribution for the GCS subscores is likely the more fundamental solution and is left to future work.

*Supplementary Methods S4 · Revision of the state names*

The names first drafted by the analytic team were "relatively stable", "mild neurological impairment", "neurological impairment with multi-organ involvement" and "respiratory-support-dependent (severe)". These were revised before blinded expert review, for two reasons.

First, "multi-organ involvement" went beyond what the data support. The variable set includes renal indices only — creatinine and blood urea nitrogen — and contains no measure of hepatic function, coagulation or other organ systems; renal replacement therapy was used in 1.4% of windows in that state.

Second, severity terms ("mild", "severe") and temporal judgments ("stable") presuppose the conclusion the outcome analysis is meant to test. A state describes a single 6-hour window, so "stable" imports a judgment about time; "mild" and "severe" import a risk ordering, and the naming experts were deliberately shown no outcome data precisely so that naming would not be shaped by outcome.

The four revised names share a two-part structure: neurological status first, then the organ-function or support feature that best distinguishes that state from the other three. The final labels were then evaluated in a blinded review by a critical care nurse scientist (Y.Z., RN, PhD), who saw the state profiles but no outcome data.

*Supplementary Methods S5 · Composition of the missing data*

Window-level missingness for laboratory variables ranges from 62% to 71%, but nearly every patient has measured values at some point during the ICU stay. For creatinine, 32.7% of windows carry a measured value, 42.8% are carried forward from a measurement made within the preceding 12 h in the same patient, 24.5% take the training-set median, and only 1.9% of windows belong to patients with no creatinine measurement at all. Between 96.9% and 97.3% of patients have at least one measurement of each laboratory variable, with a median of 3 measurements per patient.

Forward filling within 12 h is defensible for these variables because they change slowly relative to the window length. The variables that change fast enough to make forward filling inappropriate — lactate, pH, arterial blood gases — are exactly those excluded by the 70% missingness rule.

*Supplementary Methods S6 · Age de-identification*

MIMIC-IV records anchor_age as 91 for all patients aged 90 or older, so the field contains no values between 90 and 91. Age at admission is derived as anchor_age plus the difference between admission year and anchor_year; this derived value is not itself capped and reaches 101 in this cohort, with 73 patients above 91 years.

The primary model caps age at 90 to keep an artefact of de-identification out of the covariate. Refitting every model with the uncapped derived age changed the state odds ratios by at most 0.008 in absolute terms.

*Supplementary Methods S7 · Temporal reproducibility across admission eras*

MIMIC-IV date-shifts admission times per patient, so the recorded admission year is not the true year. The `anchor_year_group` field is the only usable temporal signal, and it divides this cohort into five roughly equal eras of 1150 to 1398 patients.

Practice changed materially over the period covered. ICU mortality fell from 13.8% among patients admitted in 2008–2010 to 7.6% in 2020–2022, so admission era is a real contrast rather than a null one.

**Mapping.** Applying the frozen primary model to each era, state prevalence tracked that secular change. Neurologically preserved-low support rose from 57.7% to 69.4% of windows across the five eras; neurological impairment-respiratory support fell from 19.3% to 11.4%; neurological impairment-renal dysfunction fell from 15.6% to 8.9%; neurological impairment-low support moved from 7.4% to 10.4%.

Self-transition probabilities were far more stable than prevalence: 93.5–96.8% for neurologically preserved-low support, 73.5–81.4% for neurological impairment-low support, 83.8–86.7% for neurological impairment-renal dysfunction, and 82.1–87.6% for neurological impairment-respiratory support.

**Refitting.** The model was fitted again from scratch within an early era (2008–2013, 2500 patients, 23063 windows) and a late era (2017–2022, 2550 patients, 26043 windows). The middle era was left out of this contrast so that the two periods are separated in time. Both refits recovered a four-state structure, with restart stability of 0.936 and 0.959. Agreement with the primary assignment was ARI 0.860 in the early era and 0.899 in the late era; pooling both refits gave ARI 0.879.

This is the strongest reproducibility evidence available within a single database, and it is not external validation. One institution across time is not another institution.

*Supplementary Methods S8 · No forward fill*

The plan specified a sensitivity analysis repeating the work with no forward filling at all, every unmeasured window taking the training-set median.

With the full feature set this specification is not estimable under the present emission model. Without forward filling, each of the eight laboratory variables takes a single value in 66–71% of windows, and a diagonal-covariance normal emission cannot be fitted to a variable that is mostly a point mass: expectation-maximization drives the per-state variance to zero. None of 50 restarts converged, at any variance floor tested between 0.05 and 0.5. This is the same failure as robust standardization (S3), and it has the same cause: a continuous emission asked to model what imputation has turned into a near-discrete variable.

Removing the eight laboratory variables removes the point masses. The specification then converges with a restart stability of 0.899, agreement with the primary model of ARI 0.629, and a mortality gradient across the four recovered states from 2.8% to 33.7%.

The question the analysis was designed to address — whether the four states are an artefact of imputation — is answered more directly by the prespecified analysis that removes the laboratory variables outright while retaining forward filling (ARI 0.754, Section 3.6).

*Supplementary Table S1 · Variable to itemid mapping*

See the accompanying file supplementary_table_s1_itemid_mapping_EN.csv.

*Supplementary Figures*

**Supplementary Fig. S1.** Full-resolution state evolution pathways across all twelve 6-hour windows in the full cohort (n=6368). Vertical ordering of states is fixed across columns to make transitions readable.

**Supplementary Fig. S2.** Clinical feature heatmap across the four dynamic states. Cell values are raw medians; color encodes the z-score of each variable across the four states.

**Supplementary Fig. S3.** Silhouette coefficients for the four-state assignment, computed over the full training set. Silhouette is reported as a descriptive measure of static geometric separation in the raw feature space. It was not a state-selection criterion: it evaluates each window as an independent point and is therefore blind to the transition structure that distinguishes these states from clusters.
