## Supplementary material for "Dynamic Clinical States and Transitions During the First 72 Hours of Intensive Care After Acute Stroke": Statistical Analysis Plan

**Prespecified Modeling Strategy and Statistical Analysis Plan**

**Study:** Dynamic clinical states and their evolution during the early ICU course of acute stroke (MIMIC-IV v3.1)

*0. Status of this document*

This plan formalizes the analysis-relevant content of the study design document written before any model was fitted. It is a restatement, not a reconstruction: every item in Sections 1–11 is taken from that document, and the section numbers of the source are given so each item can be checked against it. The original design document is available on request.

This plan was not registered with an external registry and carries no third-party timestamp. Readers should weight it accordingly. What it does establish is which criteria were chosen before the results were seen, which matters most for the choice of the number of states, where a criterion selected after seeing the fit would carry little weight.

Section 12 lists every departure from the plan, and Section 13 lists prespecified analyses that were not performed. Both are stated in full.

*1. Objectives*

Primary: identify a small number of interpretable, recurring clinical states from longitudinal ICU data in acute stroke, and characterize the transitions between them.

Secondary: assess whether the state a patient currently occupies is associated with subsequent organ-support escalation and death; compare state distributions across stroke subtypes.

Explicitly deferred to future work: prediction of the next state (design document, stage four).

*2. Population (source §4.1–4.3, §6.1–6.5)*

Adults with an acute stroke code (ischemic stroke, intracerebral hemorrhage, subarachnoid hemorrhage) among the first three diagnosis positions of a hospital admission that included an ICU stay. Admissions also coded for traumatic brain injury were excluded. Only the first qualifying ICU stay per patient was retained. Stroke was identified from ICD-9 and ICD-10 codes; subtype was retained as a stratification variable throughout.

*3. Observation period and time windows (source §5.1–5.3, §7.1–7.4)*

Time zero is ICU admission. Observation continues to ICU discharge, ICU death, or the end of the analysis period, whichever is earliest. The ICU course is divided into consecutive, non-overlapping 6-hour windows.

The 6-hour window was chosen in advance as a balance between sensitivity to clinical change and data completeness: shorter windows generate missingness and noise, longer windows conceal acute change, and 6 hours matches ICU monitoring practice and short-horizon prediction.

*4. Variables (source §6.1–6.2, §10.1–10.7, §12.1–12.3)*

Variables were organized in advance by physiological domain: neurological, respiratory, circulatory, renal-metabolic, hematological, inflammatory, hepatic and other organ function, plus treatment variables. Continuous variables were to be summarized within each window; treatment variables were to be coded as present when overlapping the window.

*5. Operational definition of a dynamic clinical state (source §14, paragraphs on definition)*

A dynamic clinical state is the multidimensional clinical phenotype of a patient within a fixed time window, constituted jointly by neurological function, respiratory, circulatory and metabolic status, organ function, and treatment intensity.

States are not defined in advance by death, intubation or a SOFA threshold. They are identified from the data by an unsupervised or probabilistic model. A state recovered by the model was required, in advance, to satisfy all of the following:

- be statistically distinguishable from the other states;
- be clinically interpretable;
- be reproducible across data subsets;
- show a plausible association with subsequent clinical events;
- not be completely dominated by any single variable.

*6. Missing data (source §8.1–8.2, §13.1–13.4)*

Variables with window-level missingness above 70% were not to enter the primary state model. Clinically important variables with high missingness could be carried into sensitivity analyses. Missingness alone was not to be grounds for dropping key neurological variables.

Imputation: limited forward fill within patient, up to 6 h for vital signs and up to 12 h for laboratory values; beyond that horizon the value is missing. Remaining missing values are filled with the training-set median. Missingness indicators were to be added for important variables.

Future window values must never be used to impute past windows.

*7. Standardization and data splitting (source §7.3, §14.1)*

Continuous variables are standardized to z-scores using training-set means and standard deviations. Standardization parameters must not be computed on the full dataset. Patients are split 70/30 into training and test sets at the patient level, with all windows of a patient in the same set.

*8. State identification (source §16.1–16.4)*

**Model class.** A latent-state model with time-dependent structure was specified as the primary method — hidden Markov models and their relatives — rather than clustering. The reason was recorded in advance (§16.3): conventional clustering treats each window as independent and cannot naturally represent state ordering, state duration, transition probability, or correlation between adjacent windows. Clustering was to serve as a benchmark, not as the sole primary method.

**Candidate number of states.** Models with 3, 4, 5 and 6 states were to be fitted. The design document states that more than 6 was not advisable for a first analysis.

**Evaluation criteria, fixed before fitting.** Candidate models were to be judged on:

| **#** | **Criterion** |
| --- | --- |
| 1 | Statistical fit (Bayesian information criterion; log-likelihood) |
| 2 | Share of patients and windows in each state |
| 3 | Whether any state is very small |
| 4 | Whether states differ clinically in a substantive way |
| 5 | Whether results are stable across random initializations |
| 6 | Whether clinicians can reasonably interpret the states |

**The decisive rule, also fixed in advance.** The number of states must not be chosen on the Bayesian information criterion alone. The stated reason: in latent class trajectory analysis, small changes in model specification can materially alter both the recovered classes and their apparent clinical associations, so stability and clinical interpretation must enter the decision. The design document states explicitly that the final number of states is not determined by statistical indices alone.

This is a set of evaluation dimensions with a stated exclusion, not a deterministic decision rule. No weighting between criteria was fixed in advance, and the final choice was a judgment across all six.

**Small-state rule.** A state occupying below approximately 1–2% of windows was to trigger reconsideration of the number of states.

*9. State naming (source §16.5, §17.2)*

States are identified by the model first and named afterwards by clinicians from the variable profile. Naming does not use outcome information.

*10. Transitions (source §12.1–12.3, §18.1–18.4)*

Transition probabilities between adjacent windows; state duration in windows, with median duration, self-transition probability and time of first appearance; and the five to ten most common state paths, displayed as a Sankey or state-transition diagram.

*11. Outcomes and association analysis (source §13.1–13.3, §22.1–22.6)*

**Construct validity.** Compare GCS, mechanical ventilation rate, vasopressor rate, CRRT rate, treatment intensity and ICU mortality across states.

**Predictive validity.** Relate the current state to mechanical ventilation within 12 h, vasopressor initiation within 12 h, ICU death, and live ICU discharge.

**Adjustment.** Age, sex, stroke subtype, Charlson comorbidity index. Effect sizes with 95% confidence intervals are to be reported, not p values alone.

**Stability.** Bootstrap, different random seeds, different sample subsets, different window lengths, different numbers of states, different missing-data strategies.

**Temporal reproducibility.** Fit or map states separately in earlier and later admission years and compare state profiles, state proportions, transition probabilities and outcome associations.

*12. Deviations from the plan*

Each deviation below was made during execution, with the reason recorded at the time.

| **Item** | **Plan** | **Executed** | **Reason** |
| --- | --- | --- | --- |
| Observation period | The design document is internally inconsistent: §7.2 specifies the first 7 ICU days, while the cohort-construction section specifies extraction of the first 72 h | First 72 h | The 72 h specification was followed. The analysis is framed throughout as the early ICU course, and the inconsistency is disclosed here rather than resolved silently |
| Minimum ICU stay | ≥24 h | ≥12 h | A 24 h threshold systematically excludes patients who die within the first ICU day, precisely the most severely ill. The 24 h threshold is retained as a sensitivity analysis (ARI 0.946) |
| Candidate states | 3–10 in §16.4; 3–6 elsewhere in the same document | 3–6 | The document itself advises against more than 6 for a first analysis |
| Association model | Logistic, mixed-effects logistic, or Cox regression | Generalized estimating equations, exchangeable working correlation, clustered by patient | Windows are repeated observations within patient; GEE gives a population-averaged estimate with a correct variance under that clustering. For ICU death each window is treated as a discrete-time hazard interval |
| Missingness indicators | To be added for important variables in the primary model | Moved to a prespecified sensitivity analysis (ARI 0.955) | Panel-level indicators encode local ordering behavior. Keeping them out of the primary model keeps the state definition transportable to other ICU databases, where ordering practice differs |
| Standardization | Z-score (§7.3) | Z-score | A robust median/IQR alternative was added after the plan and prespecified before fitting; it degenerated to a single-state solution under this emission model and could not be used |
| GCS verbal | Not addressed in the plan | Excluded from the primary model; three alternative handlings carried as sensitivity analyses | Verbal response is unassessable in intubated patients (29.0% of windows missing, 80.2% of those ventilated). Any single imputation biases the neurological dimension in a known direction |
| Sparse-cell reporting | Not addressed in the plan | Odds ratios reported only for states contributing ≥10 outcome events | Fixed before the affected cells were examined |

*13. Prespecified analyses: status*

Section 26 of the design document lists fifteen sensitivity analyses under the heading "at least the following". A subset was carried out. The full list is reproduced below with the status of each, together with the prespecified items from other sections, so that what was planned and not done is visible rather than inferred.

The executed subset was chosen to cover the dimensions on which the state structure could plausibly be an artefact — imputation, treatment variables, the variable set, cohort restriction, window length and within-window aggregation — rather than to exhaust the list. That is a judgment made during execution and is recorded here as such.

***Performed***

| **Analysis** | **Source** | **Result** |
| --- | --- | --- |
| Refit without treatment variables | §26 | ARI 0.935 vs primary |
| Different imputation scheme: laboratory missingness indicators added | §26, §13.3 | ARI 0.955 |
| Different imputation scheme: no forward fill | §8.3, §26 | Not estimable with the full feature set (no restart converged in 50); estimable with laboratory variables removed (restart stability 0.899, ARI 0.629) |
| Refit with routinely available variables only, implemented as removing all laboratory variables | §26 | ARI 0.754; the renal dysfunction state is not recovered |
| Complete-case analysis | §26 | 8048 training windows (18.6%) with all 17 continuous variables measured; four states recovered, ARI 0.772 vs primary. The retained windows are neither contiguous nor a random subset |
| Exclude patients with high sedation depth | §26 | 931 patients (14.6%) excluded; ARI 0.965 vs primary. Sedation depth proxied by sustained sedative infusion, as RASS was not extracted |
| Ischemic stroke only | §26 | ARI 0.895 |
| Different window length: 12 hours | §7.5, §26 | ARI 0.824 |
| Within-window mean instead of last value | §12 | ARI 0.875 |
| Different state counts (K = 3–6) | §26, §16.4 | Reported in Supplementary Methods S2 |
| Restore the 24 h minimum stay | §9 | ARI 0.946 |
| Temporal reproducibility across admission years | §14.3, §22.5 | Frozen model mapped onto five eras; refitted within an early and a late era (ARI 0.860 and 0.899) |

***Not performed***

| **Analysis** | **Source** | **Note** |
| --- | --- | --- |
| 4-hour and 8-hour windows | §7.5, §26 | 12-hour windows were tested; the shorter windows were not |
| Observation period of 3 and 14 days | §26 | The analysis was fixed at 72 h throughout |
| Exclude patients dying within 24 h of ICU admission | §26 | The related 24 h minimum-stay analysis was performed instead |
| Exclude patients receiving palliative care or limitation of life support | §26 | Treatment-limitation decisions were not extracted |
| Hemorrhagic stroke only | §26 | The ischemic-only analysis was performed; the hemorrhagic counterpart was not |
| Alternative latent-state models (hidden semi-Markov, switching state-space) | §16.1, §26 | Only the hidden Markov specification was fitted |
| Stratification by neurological versus general ICU | §26 | Care-unit stratification was not analyzed |
| Stratification by mechanical ventilation status | §26 | Not analyzed; ventilation enters the model as a state-defining variable |
| Bootstrap stability | §22.4 | Stability was assessed by restart stability, subset refitting, held-out reproducibility and admission-era refitting |
| Live ICU discharge as an outcome | §13.2, §22.2 | Death and live discharge are competing outcomes; only death was analyzed |
| Next-state prediction | Stage four | Stated in the manuscript as the intended next study |

*14. Sensitivity analyses actually performed*

Eleven analyses, each compared with the primary model by the adjusted Rand index: laboratory missingness indicators added; ICU stay ≥24 h; all treatment variables removed; AIS only; within-window mean instead of last value; GCS verbal imputed at the median; GCS verbal estimated by regression; GCS verbal set to the floor score; 12-hour windows; all laboratory variables removed; robust standardization.
